# TIM3 blockade with hypomethylating therapy restores NK and cytotoxic CD4+ T cell activity in patients with AML or MDS

**DOI:** 10.1101/2025.11.14.25339889

**Authors:** Jani Huuhtanen, Sofia Forstén, Brittany Ford, Johannes Smolander, Oscar Brück, Sofie Lundgren, Anna Kreutzman, Olli Dufva, Matti Kankainen, Mette Ilander, Hanna Lähteenmäki, Tiina Kasanen, Jay Klievink, Judith Leitner, Peter Steinberger, Mika Kontro, Marc Pelletier, Harri Lähdesmäki, Catherine Sabatos-Peyton, Mikael Rinne, Kimmo Porkka, Karita Peltonen, Satu Mustjoki

## Abstract

Occasional complete responses to immune checkpoint inhibitor therapies suggest that acute myeloid leukemia (AML) and myelodysplastic syndrome (MDS) are immune-sensitive when appropriately targeted. Here, we analyzed AML/MDS patients treated with anti-TIM3 sabatolimab and decitabine in a phase Ib clinical trial (NCT03066648) using single-cell RNA and T cell receptor (TCR) sequencing and functional co-culture assays. Unlike T cell restricted *CTLA4* and *PD1*, *TIM3* was broadly expressed across natural killer (NK), myeloid, and T cell populations. Therapy induced expansion of cytotoxic NK cell subsets and enhanced type I interferon signaling. Less than 1% of bone marrow CD8+ T cells showed canonical exhaustion phenotypes, and the treatment preferably expanded small CD8+ T cell clones in responders. Responders had more cytotoxic CD4+ T and B cells which was exemplified by a patient with an outstanding complete response of 23-months. Over >20% of this patient’s lymphocytes were CD4+ T-cell large granular lymphocyte leukemia (T-LGLL) cells that expressed a TCR capable of recognizing autologous blasts.

**Significance:** Anti-TIM3+decitabine offers a distinct mechanism compared to anti-PD1 and anti-CTLA4: it preferentially enhances innate and unconventional cytotoxic lymphocyte populations. Furthermore, our findings suggest that clonal expansion of self-antigen–specific T cells, such as T-LGLL, may augment anti-leukemic immunity in the context of immunotherapy.

## Introduction

Acute myeloid leukemia (AML) and myelodysplastic syndrome (MDS) are cancers of the hematopoietic stem cells (HSCs), caused by differentiation arrest and accumulation of precursor hematopoietic cells. The success of allogeneic hematopoietic stem cell transplantation (allo-HSCT) supports the idea that immunotherapy can have curative potential in AML/MDS. Allo-HSCT is associated with life-threatening toxicities and requires donor availability, thereby limiting its use to certain patient populations and contributing to the search for more precise novel immunotherapies.

It remains unclear why currently available immune checkpoint inhibitors, anti-CTLA4, anti-PD(L)1, and anti-LAG3, have shown only modest efficacy in patients with AML/MDS even when combined with standard-of-care hypomethylating agents (HMAs, i.e., decitabine or azacytidine)^1,2^. Previous correlative studies of anti-CTLA4 or anti-PD1 in combination with HMAs have shown only little effect on the reversal of T cell exhaustion^1–3^. Interestingly, T cell activation has been linked to immune-related adverse-effects such as hepatotoxicity rather than therapeutic benefit^3^. The lack of efficacy might be explained by a relative low rate of exhausted T cells^4^ and the exceptionally low tumor mutation burden (TMB) of AML/MDS^5^, both of which stand in stark contrast to melanoma and non-small lung cancer, where high TMB has been linked to greater benefit from immune checkpoint inhibitor therapies^6^. This might suggest that spontaneous endogenous responses against malignant HSCs are scarce, necessitating outsourced immunity from cellular therapies. However, myeloid malignancies are among hematological cancers which are most sensitive to natural killer (NK) cell killing^7^, and autoimmunity against HSCs driven by (NK-like) T and NK cells is commonly seen in immune aplastic anemia^8^ and T cell large granular leukemia (T-LGLL)^9^, highlighting immune effector pathways that may not be fully engaged by conventional checkpoint inhibitors such as anti-CTLA4 or anti-PD1.

Among immune checkpoint receptors, TIM3 is an attractive target, as it is widely expressed by immune cells, including NK cells, dendritic cells (DC), monocytes, and macrophages, but also by terminally exhausted CD8+ T cells, T helper 1 CD4+ T cells, and regulatory T cells (T_REGS_)^10^. In AML/MDS, TIM3 is of significant interest as it is also expressed by malignant myeloid cells^10^, including leukemic stem cells^11,12^ that might promote relapse following conventional chemotherapies. The combination of anti-TIM3 with HMAs has shown initial response rates up to 50-60% in patients with AML^13^ or MDS^14,15^ in phase I/II trials, but the mechanism of action is incompletely understood^16^.

Here, we conducted comprehensive immunomonitoring of the phase Ib trial (NCT03066648)^16^ evaluating decitabine in combination with the anti-TIM3 antibody sabatolimab (MBG453). We studied paired bone marrow (BM) and peripheral blood (PB) samples from 11 unfit for chemotherapy and newly diagnosed (ND) or relapsed/refractory (R/R) AML patients and 1 MDS patient refractory to HMA with scRNA+T-cell receptor (TCR)αβ-seq and flow cytometry and performed co-culture assays of primary immune and leukemic cells with scRNA+TCRαβ-seq readout in a patient with a durable complete response (CR). Additionally, we compared our scRNA+TCRαβ-seq data to 160 BM samples from healthy donors and patients with AML and 9 other hematological malignancies. Unlike *PDCD1* (encoding PD1) and *CTLA4* that were restricted to T cells, *HAVCR2* (TIM3) was expressed on a broad range of NK, myeloid, and leukemic cells. In concordance, our results show that the effects of TIM3 blockade *in vivo* are mainly observed in these cell types and diverge from those seen upon treatment with anti-PD1 and anti-CTLA4, helping to position this therapy in the growing immuno-oncology arsenal.

## Results

### Anti-TIM3 sabatolimab in combination with HMA decitabine

We enrolled 12 patients with newly diagnosed (ND) or relapsed/refractory (R/R) AML unfit to intensive chemotherapy (*n*=11) and 1 patient with MDS to the phase I trial (**Supplementary Data 1**). The treatment started with 1 priming cycle of decitabine monotherapy (20 mg/m2 on days 1-7) followed by treatment using a combination of decitabine and sabatolimab (3-10 mg/m2 on day 8). We routinely collected PB samples from screening (SCR), following the priming cycle of decitabine (C1D8), and then on the first day of every cycle, and BM aspirate samples from screening, on the first day of every third cycle and at the end of treatment (EOT, **Fig. 1A**).

**Figure 1:**
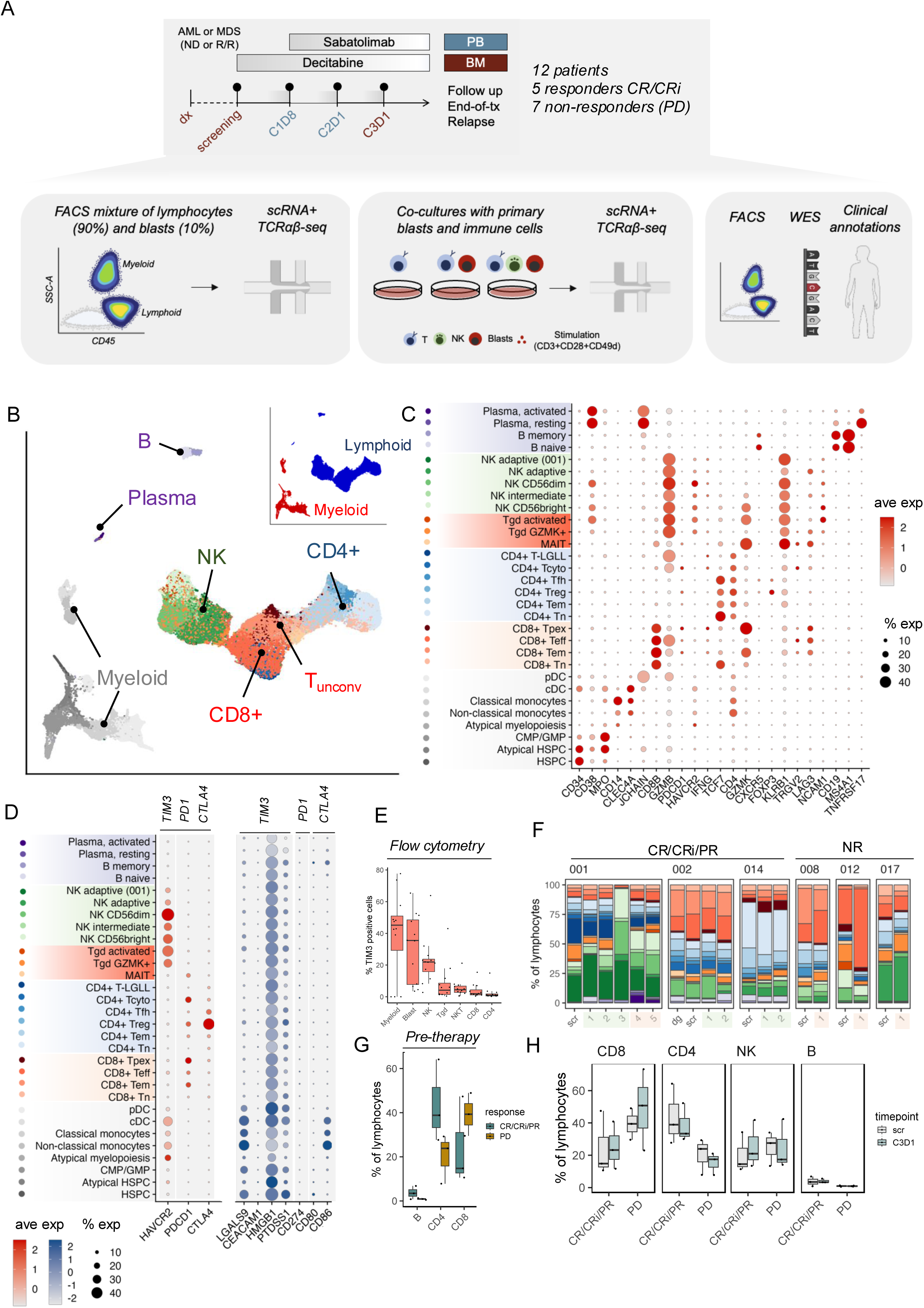
Immune responses to anti-TIM3+HMA treatment in AML/MDS. (A) Study schematic illustrating patient treatment (sabatolimab, decitabine), sample acquisition (PB: peripheral blood; BM: bone marrow), and multi-omic analyses (flow cytometry (FACS), scRNA+TCRαβ-seq, co-cultures, whole-exome sequencing WES). (B) UMAP of bone marrow scRNA-seq profiles from AML/MDS patients (n=7 patients, 20 samples) colored by manually annotated immune cell lineages. (C-D) The average expression (color) and percentage of cells expressing (dot size) of canonical markers used for cluster annotation (C) or key immune checkpoint receptors and their main ligands (D). (E) The proportion of TIM3-expressing bone marrow cells in pre-treatment samples analyzed with flow cytometry within their respective category (*n*=12 patients). (F) The proportion of lymphocyte populations in the bone marrow across different time points analyzed with scRNA-seq. (G-H) The proportion of main lymphocyte populations in the bone marrow in pre-treatment samples (G) and through treatment (H) (responders: CR/CRi/PR *n*=3; non-responders: NR *n*=4).

In AML, the combination treatment resulted in responses for 5 patients: 3 complete responses (CR) and 2 complete responses with incomplete hematological recovery (CRi). The other 8 patients had no clinically meaningful benefit (progressive disease, PD). The patient with MDS (017) was azacytidine-refractory and thus treated with sabatolimab monotherapy, but did not gain a clinical response and had PD.

### Single-cell characterization of immune and leukemic cells in AML/MDS BM

We observed that prior immunological characterizations of AML/MDS have been limited by insufficient sampling of the immune cell repertoire^2,4,17^, largely due to the predominance of malignant myeloid cells in BM aspirates, which cannot be reliably separated from non-malignant cells without genetic information^18–20^. Given that the proposed mechanism of action for anti-TIM3+HMA involves reinvigoration of lymphocytes, we employed a strategy to enrich for lymphocytes by single-cell profiling a cell mixture composed of 90% lymphocytes and 10% myeloid cells, including the putative leukemic blast cells (**Fig. 1A, Supplementary Fig. 1A**).

After quality control, we had 112,449 BM scRNA+TCRαβ-seq profiles (*n*=7; AML *n*=6, MDS *n*=1; 20 samples). We defined cell types by first identifying broader cell lineages, including hematopoietic stem and progenitor cells (HSPC), myeloid cells, CD8+ T cells, CD4+ T cells, NK cells, unconventional T cells, and B cells, and then performed iterative clustering on each of these classes to balance over- and underfitting the data (**Fig. 1B-D**, **Supplementary Fig. 1B**).

With this strategy, we annotated 32 different cell states, including 8 different HSPC and myeloid cell, 4 CD8+ T cell, 6 CD4+ T cell, 5 NK cell, 3 unconventional T cell, and 4 B cell phenotypes (**Fig. 1B-D, Supplementary Fig. 2A-B**). This aligns with previous AML scRNA+TCRαβ-seq studies^2–4,17^, but due to our enrichment strategy, we were able to provide a more detailed annotation of lymphocytes, especially NK cells^7^.

The largest diversity of immune cell states was seen in patient 001, who achieved an outstanding response in our cohort (>2 years in CR), harboring three unique cell populations: cytotoxic CD4+ T-LGLL cells, patient-specific adaptive NK cells, and activated plasma cells (**Fig. 1B-C, Supplementary Fig. 2A**). Patient 001, was a female in their 70s, presenting with therapy-related AML with mutations in *TET2*, *SRSF2*, *RUNX1*, and deletion 7 (del7), and the patient was deemed unfit for conventional chemotherapy. Patient 001 had been previously treated for breast cancer with postoperative radiotherapy (50 Gy) in 2006 and, intriguingly, diagnosed with CD4+ T-LGLL in 2012, which required no treatment. T-LGLL is an autoimmune-like manifestation that results in indolent clonal outgrowth of mature T cell clones^21^. Remarkably, the patient achieved CR after the first cycle and achieved minimal residual disease (MRD) negativity before cycle 6 day 1 (C6D1) as the best response (more detailed history in **Supplementary Note 1**). The patient relapsed after 2 years of durable remission.

### TIM3 is expressed on myeloid cells, NK cells, and CD8+ T cells especially in AML

In the BM cells, the highest *HAVCR2* (encoding TIM3) expression was seen in NK cells (immature CD56^bright^, intermediate NK cells, mature CD56^dim^, adaptive NK cells), myeloid cells (conventional DCs, classical and non-classical monocytes, atypical myelopoiesis), and unconventional T cells (activated gamma-delta T cells (T_GD_), GZMK positive Tgd^2^) (**Fig. 1D**). Besides high expression in these subsets, *HAVCR2*/TIM3 was also expressed at lower levels in CD8+ T cells (effector T cells (T_EFF_)), pre-exhausted T cells (T_PEXH_), T_REGS_, and HSPCs. The RNA expression was also confirmed with elevated protein level expression by flow cytometry (**Fig. 1E**). The expression profile of *HAVCR2*/TIM3 was in stark contrast to *PDCD1* (encoding PD1) and *CTLA4*, that are the previously targeted immune checkpoint receptors in AML/MDS. *PDCD1* expression in AML/MDS BM was restricted to CD8+ T cells (T_PEXH_, T_EFF_, T_EM_), CD4+ cells (cytotoxic CD4+ T cells, T_REG_), and mucosal-associated invariant T cells (MAIT). *CTLA4* was expressed mostly on CD4+ T cells (CD4+ T_REG_, CD4+ follicular helper T cells (CD4+ T_FH_)).

Conversely, the putative TIM3 ligands (*LGALS9, CEACAM1, HMGB1,* and *PTDSS1*)^22^ were more highly expressed on putative blast cells compared to the ligands for PD1 (*CD274*, encoding PDL1) and CTLA4 (*CD80* and *CD86*), which showed only minimal expression (**Fig. 1D**). In previous preclinical study, TIM3 ligand expression on the target leukemia cells was shown to be a biomarker for response to TIM3-blockade^23^ - in our study, all patients had abundant TIM3 ligand expression in myeloid cells as well as *HAVCR2*/TIM3 expression in lymphocytes (**Supplementary Fig. 3A-B**),

We verified these findings by integrating published scRNA-seq samples from a range of hematological malignancies (AML^17,18^ *n*=13, B-cell acute lymphoblastic leukemia [B-ALL]^24–26^ *n*=19, T-cell ALL [T-ALL]^26^ *n*=2, monoclonal gammopathy of unknown significance [MGUS]^33^ *n*=5, multiple myeloma [MM]^27^ *n*=7, smoldering MM [SMM]^27^ *n*=11, and healthy controls [HC]^18,27,28^ *n*=32). This pan-heme scRNA-seq analysis of over 500,000 cells confirmed our finding that *HAVCR2*/TIM3 was expressed at the highest level in NK and myeloid cell subsets (**Supplementary Fig. 4A**). In HSPC populations, AML patients had generally upregulated *HAVCR2*/TIM3 expression compared to healthy controls (**Supplementary Fig. 4B**).

### Responding patients have increased amounts of CD4+ T cells and B cells in their BM

Interestingly, in the scRNA+TCRαβ-seq data, the responding patients had more CD4+ T cell and B cell lymphocytes in their BM prior to treatment than non-responding patients (**Fig. 1F-H, Supplementary Fig. 2B**). On the contrary, the non-responding patients had a higher proportion of cytotoxic CD8+ T cells at baseline and post-therapy (**Fig. 1F-H**). This was also validated with the flow cytometry analysis, where especially the number of CD8+ T_EMRA_ cells was higher in non-responders than in responders in both PB and BM samples (**Supplementary Fig. 5A-C**). This is in contrast to patients who respond to anti-PD1 therapy in solid tumors^29^, suggesting that anti-TIM3 combined with HMA may act through a different mechanism.

### Anti-TIM3+HMA expands mature NK cells resulting in type I and II IFN activation in responding patients

We first focused on NK cells, given their notably high *HAVCR2* (TIM3) expression and marked expansion during successful therapy. After re-clustering the NK cells, we identified 5 different NK cell populations: cytokine-producing immature CD56^bright^ NK cells (*NCAM1*/CD56, *GZMK*, *XCL1*), intermediate NK cells^30^ (*AREG*, *PIK3R1*), cytotoxic CD56^dim^ NK cells (*FCGR3A*/CD16, *GZMB*, *PRF1*), and two populations of memory-like adaptive NK cells (*KLRC2*/NKG2C, *LAG3*) (**Fig. 2A-B**, differentially expressed genes [DEGs] in **Supplementary Data 2**, details of re-clustering in **Methods**). These NK cell states aligned with previous scRNA-seq classifications of NK cell phenotypes in the BM^7,8,31–33^, and the known maturation trajectory was supported by pseudotime analysis^34^ (**Fig. 2C**). Adaptive NK cells, which are associated with CMV infection^35^, were seen in every scRNA+TCRαβ-seq profiled patient in concordance with noted CMV seropositivity (clinical details in **Supplementary Data 1**). Patient 001 with a concomitant CD4+ T-LGLL diagnosis had a unique subset of adaptive NK cells, which expressed *CD3D* transcript^36^ and lower levels of *LAG3* than the traditional adaptive NK cells but did not express TCR, excluding them from being T cells. This population clustered together with other adaptive NK cells in our pan-hematological malignancy cohort, strengthening the hypothesis that these are adaptive NK cells and not other innate lymphocyte cells (**Supplementary Fig. 6A-D**).

**Figure 2:**
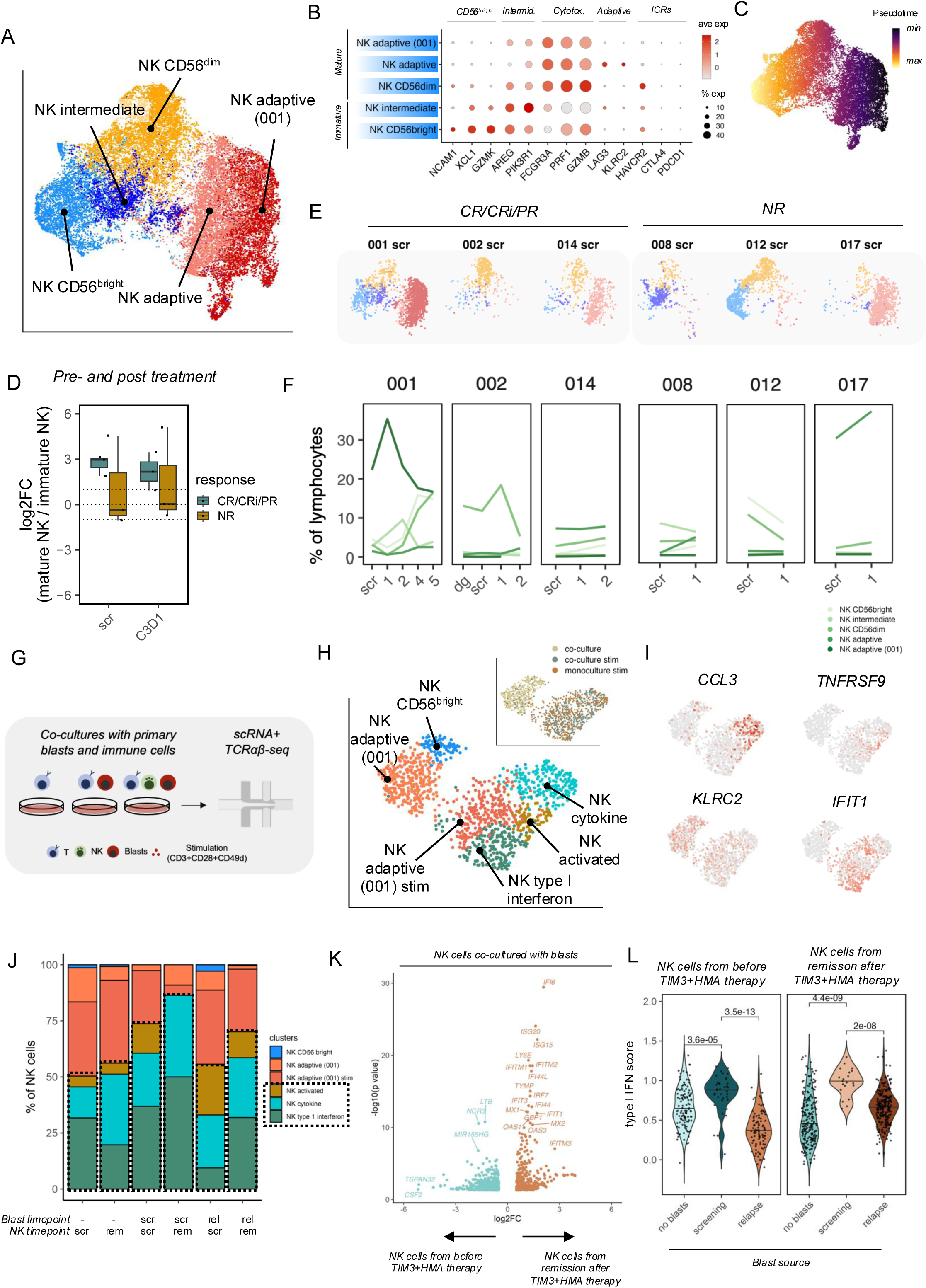
NK cell responses to anti-TIM3+HMA treatment. (A) UMAP of bone marrow NK cells from AML/MDS scRNA-seq profiles (*n*=7 patients, 20 samples) colored by manually annotated NK cell subtypes. (B) Average expression (color) and percentage expression (dot size) of canonical marker genes for NK cell subtype annotation. (C) NK cells colored by pseudotime analysis, indicating developmental trajectory. (D) The ratio of mature (CD56^dim^ and adaptive) versus immature (CD56^bright^ and intermediate) NK cell abundance in responders (CR/CRi/PR) and non-responders (NR) at baseline (C0D1) and C2D1. (E) Proportions of NK cell subtypes within total lymphocytes per patient, grouped by clinical response (CR/CRi/PR vs. NR) from scRNA-seq data. (F) Percentage of total NK cells within lymphocytes over treatment time points (scr, 1, 2) for individual patients. (G) Experimental co-culture design with primary patient blasts and immune cells from patient 001, with or without antibody-based T cell stimulation with scRNA+TCRαβ-seq readouts. (H) UMAP of NK cells from patient 001 CD8+ T cells when co-cultured with blasts and MNCs with and without T cell stimulations. (I) Expression of selected phenotype markers in NK cells. (J) Proportions of NK cell subtypes under different co-culture conditions with blasts and MNCs from screening or remission samples as measured by scRNA-seq. (K) Differentially expressed genes in NK cells from patient 001 co-cultured with autologous blasts before and at remission, after TIM3+HMA therapy. (L) Type I interferon score distributions in NK cells from patient 001 before and after TIM3+HMA therapy when co-cultured with blasts from before and after TIM3+HMA therapy or without blasts as measured by scRNA-seq.

The effects of anti-TIM3+HMA treatment in patients’ NK cells were heterogenous. In general, responding patients had more mature NK cells (CD56^dim^ and adaptive NK cells) than immature populations (CD56^bright^ and intermediate) (**Fig. 2D**). The responding patient 002 had a dominating CD56^dim^ population, whereas responding patients 001 and 014 had dominating adaptive NK cell populations (**Fig. 2E)**. Although the proportion of NK cells from lymphocytes increased in responders (**Fig. 1H**), we did not observe an increase in a specific NK cell population. The most drastic response to therapy was seen early in responding patients 001 and 002, where the mature adaptive and CD56^dim^ NK cell populations expanded almost two-fold in cycle 2 day 1 and cycle 3 day 1, respectively (**Fig. 2F**).

In responding patients, the most significant transcriptional alterations in expanded CD56^dim^ and adaptive NK cells were related to type I and II IFN pathways and NF-kB signalling (**Supplementary Fig. 7A**). This was noted by upregulation of genes related to NK cell activation (*GZMA, CD38, SLAMF7*), JAK/STAT (*STAT1, STAT4, SOCS2*), and genes related to antigen processing and presentation (*TAP1*, proteasomes *PSMB8, PSMB9, PSMB10,* HLA class I molecules). These changes were not noted in the non-responding patients, with the exception of patient 017.

Given the remarkable response in patient 001, we chose to perform functional co-culture assays^8^ of this patient’s immune cells together with the blast cells throughout therapy, including pre-treatment, treatment-induced remission, and relapse, which resulted in 17 different conditions (**Fig. 1A**, all conditions **Supplementary Data 1**).

First, to elucidate the complex interactions between NK cells and blasts, we co-cultured blasts from pre-treatment and relapse samples with immune cells obtained before treatment and during treatment-induced remission (**Fig. 2G**). As in previous studies^7,8,31^, we identified three different NK cell activation states, including cell-cell contact activated NK cells (*TNFRSF9*/CD137, *TNFRSF4*/OX40, *TNF*), cytokine-secreting NK cells (*XCL1*, *XCL2*, *CCL3*, *CCL4*), and the type I IFN responding cells (*OAS1, OAS2, MX1, OX2, IFIT1, IFIT2*) (**Fig. 2H-I**). In the scRNA-seq data, we observed more NK cells in activated clusters when co-cultured with blasts from screening (**Fig.2J**). We confirmed that NK cells co-cultured with blasts exhibited an enhanced cellular response at remission after therapy compared to NK cells pre-therapy, where the top hits were in genes related to type I IFN (*OAS1, OAS2, MX1, OX2, IFIT1, IFIT2*) (**Fig. 2K)**. As type I IFN response was the most noted therapy response in our data and in previous co-cultures of NK cells with primary AML cells^37^, we modelled *in vitro* NK cell activation with type I IFN scores. Interestingly, when the NK cells were co-cultured with blasts from relapse instead of blasts from the screening phase, we noted a significant decrease in the type I IFN activity (**Fig. 2L)**. This was evident in both NK cells from pre—treatment and remission, suggesting potential immune evasion from NK cell activation in relapse samples (**Fig. 2L**).

### Oligoclonal cytotoxic CD4+ T cells are more numerous in responders

Next, we focused on the CD4+ T cells, which were more abundant in responders in pre-treatment samples (**Fig. 1G**). We identified 6 different CD4+ T cell phenotypes: naïve CD4+ T_N_ cells (*TCF7*/TCF1, *CCR7*), effector memory CD4+ T_EM_ cells (*GZMK*, *KLRB1*), follicular helper T_FH_ cells (*CXCR5*, *PDCD1*/PD1), regulatory T_REG_ cells (*FOXP3*, *IL2RA*/CD25), and cytotoxic CD4+ T cells (*GZMB*, *PRF1*, *IFNG*) (**Fig. 3A-B**).

**Figure 3:**
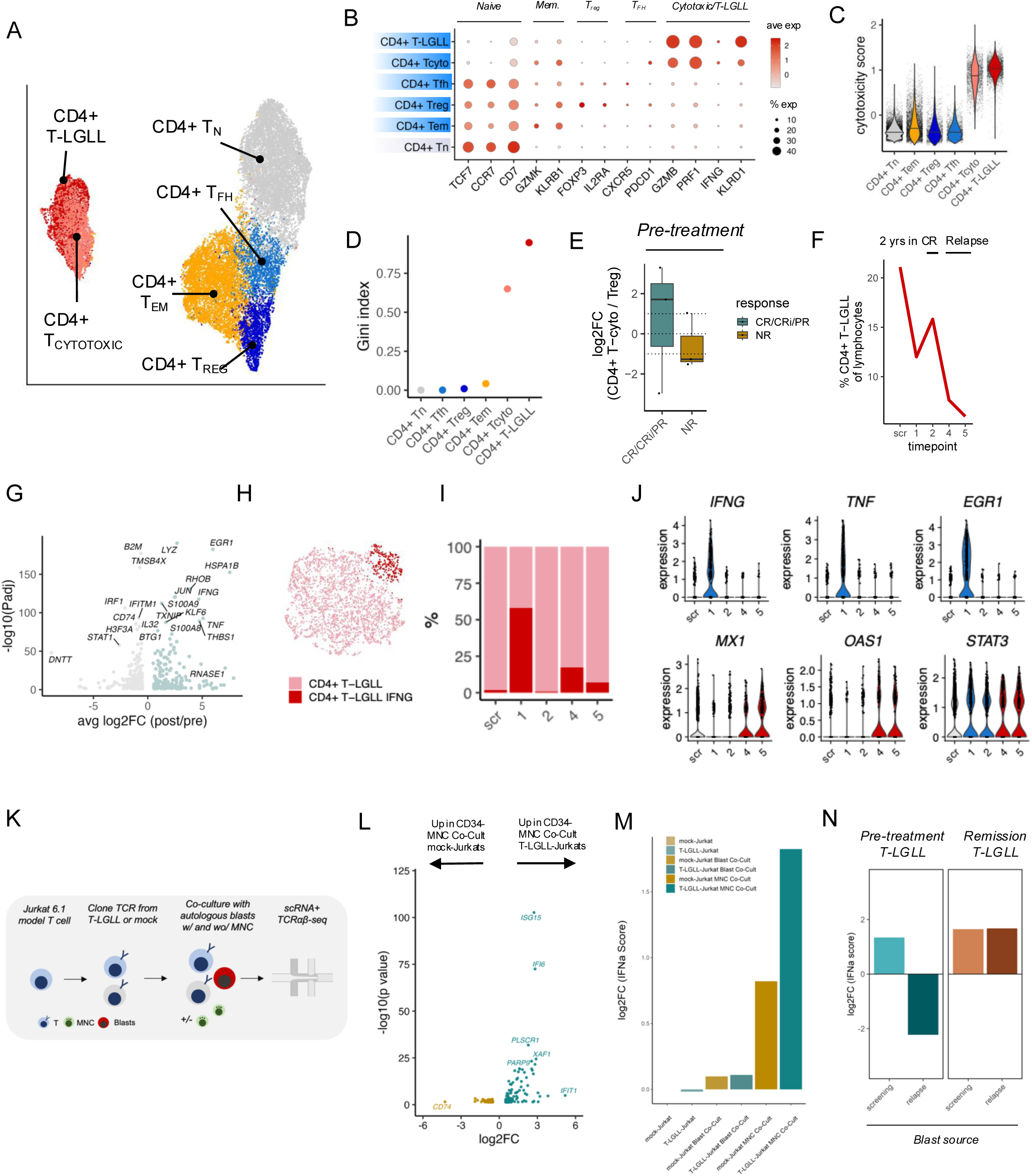
CD4+ T cell responses to anti-TIM3+HMA treatment. (A) UMAP of bone marrow CD4+ T cells from AML/MDS scRNA-seq profiles (*n*=7 patients, 20 samples) colored by manually annotated CD4+ T cell subtypes, including T-LGLL. (B) Average expression (color) and percentage expression (dot size) of canonical marker genes for CD4+ T cell subtype annotation. (C) Cytotoxicity score distribution across annotated CD4+ T cell subtypes. (D) Gini index of TCR clonality for different CD4+ T cell subtypes. (E) Log2 fold change of the ratio of cytotoxic CD4+ T cells (Tcytotoxic, T-LGLL) to regulatory T cells (Treg) in pre-treatment and post-treatment samples from responders (CR/CRi/PR) vs. non-responders (NR). (F) Percentage of the CD4+ T-LGLL clone within total lymphocytes over time for patient 001, showing periods of remission and relapse. (G) Differentially expressed genes in CD4+ T-LGLL cells before and after TIM3+HMA therapy. (H) UMAP of bone marrow CD4+ T-LGLL cells from patient 001 showing *IFNG* expressing subcluster. (I) Proportion of CD4+ T-LGLL cells in the *IFNG* subcluster at different time points in patient 001. (J) Violin plot of selected markers in CD4+ T-LGLL cells in patient 001. (K) Experimental setup for transducing Jurkat T cells with TCR cloned from T-LGLL or mock TCR, followed by co-culture with blasts and scRNA+TCRαβ-seq. The experiments did not use any T cell stimulants. (L) Differentially expressed genes in T-LGLL TCR-transduced Jurkats and control Jurkat cells after co-culture with patient MNCs. (M) Log2 fold change of IFN-α pathway score in Jurkat cells (Control or T-LGLL TCR) after co-culture with blasts or MNCs, +/− TIM3 blockade. (N) Log2 fold change of type I interferon score compared to corresponding baseline condition without co-culture in patient 001 CD4+ T-LGLL cells after co-culture without blasts from screening or remission samples.

Notably, cytotoxic CD4+ T cells and T-LGLL cells exhibited the highest cytotoxicity scores (**Fig. 3C**). Additionally, these cytotoxic CD4+ T cells showed highly increased clonality (**Fig. 3D**). The CD4+ T-LGLL cells from patient 001 manifested as an extreme pole of cytotoxic CD4+ T cells, which formed their distinct cluster with elevated expression of cytotoxic genes (*GZMB, GNLY*) and decreased exhaustion (*PDCD1*) compared to other cytotoxic CD4+ T cells (**Supplementary Fig. 8A-C**). These oligoclonal, cytotoxic CD4+ T cells were more abundant in responders, particularly relative to the ratios of immune suppressive T_REG_ cells (**Fig. 3E**). In the exceptional responder patient 001, the CD4+ T-LGLL clonotype accounted for 21.10% of the total lymphocytes in the pre-therapy sample and remained high in post-treatment remission samples (**Fig. 3F**). However, during relapse, the CD4+ T-LGLL clone diminished to 7.63% (**Fig. 3F**).

To gain more insights into the role of CD4+ T-LGLL cells, we first focused on the effects of the anti-TIM3+HMA treatment on these cells. Between post- and pre-therapy samples, the strongest DEGs included those associated with enhanced cytotoxic function of CD4+ T cells, including *IFNG*, *TNF,* and TCR-induced transcription factor *EGR1* (**Fig. 3G**). Notably, a subset within the T-LGLL clone expressing these molecules was the most prevalent in the first sample after the therapy start (C2D1, **Supplementary data 1**) when patient 001 still had a high amount (78%) of leukemic blast cells (**Fig. 3H-I**). During relapse (timepoint 4 and 5), the activated *IFNG* subset increased in proportion but upregulated type I IFN response genes like *MX1* and *OAS1*^38^ (**Fig. 3J**). *STAT3*, the major transcription factor implicated in the process of T-LGLL^21^, remained elevated throughout the therapy.

The phenotypic changes observed after therapy and at relapse suggest the possibility that the CD4+ T-LGLL clone may be reactive against the blast cells. Therefore, we engineered Jurkat 6.1. reporter T cells to express the TCRαβ from the CD4+ T-LGLL clone and conducted co-culture assays including either blast cells with the Jurkat reporter cells (henceforth T-LGLL-Jurkat) or with Jurkat mock transduced with an empty vector (henceforth mock-Jurkat) and analyzed them with a scRNA+TCRαβ-seq readout (**Fig. 3K**). Co-culture of T-LGLL-Jurkat with blasts showed increased signalling activity in T-LGLL cells, with elevated type I IFN reactivity genes including *OAS1, OAS2, MX2, IFIT1,* and *IFIT3* (**Supplementary Fig. 9A**). This was particularly evident when T-LGLL Jurkats were co-cultured with blasts and antigen-presenting cells containing mononuclear cells, highlighting that CD4+ T cells require activation from antigen-presenting cells (**Fig. 3L-M, Supplementary Fig. 9A**).

After showing that CD4+ T-LGLL cells show TCR-mediated reactivity against the blast cells, we wanted to understand whether the relapse blasts experienced immune evasion against the CD4+ T-LGLL clone. We performed co-cultures of primary CD4+ T-LGLL cells before and after anti-TIM3+HMA therapy with blasts from screening and relapse samples. Before therapy, CD4+ T-LGLL cells that were activated with CD3+CD28+CD49d stimulus showed strong type I IFN activity against blasts from the screening time point (**Fig. 3N**). This reactivity was lost when pretreatment CD4+ T-LGLL cells were co-cultured with blasts from the relapse time point, suggesting immune evasion (**Fig. 3N**). However, similarly stimulated CD4+ T-LGLL cells from post-therapy remission time point still exhibited type I IFN activity against the relapse blasts, indicating that they remained reactive to the autologous blasts. Unfortunately, we did not have sufficient CD4+ T-LGLL cells from the relapse timepoint to test in the co-culture assays.

### Exhausted CD8+ T cells are rare in AML

Next, we focused on CD8+ T cells, which are presumed to be the primary targets of the clinically available immune checkpoint inhibitors anti-PD1, anti-CTLA4, and anti-LAG3. We identified 4 distinct CD8+ T cell phenotypes, ranging from naïve (*CCR7*, TCF1/*TCF7*), to T_EM_ (*GZMK*), to cytotoxic effector (T_EFF_, *GZMB*, *PRF1*) with some NK cell receptor expression (*FCGR3A/*CD16), consistent with previous studies in AML^39^ and aplastic anemia^8^, and finally to a putatively pre-exhausted state (*PDCD1*/PD1, *LAG3*, *HAVCR2*/TIM3, **Fig. 4A-C**). As shown above, the CD8+ T cells were more abundant in non-responders than responders (**Fig. 1G**), with the predominant CD8+ phenotype being T_EFF_ cells. However, responders had higher proportion of naïve/central memory T cells compared to non-responders (**Fig. 4D**).

**Figure 4:**
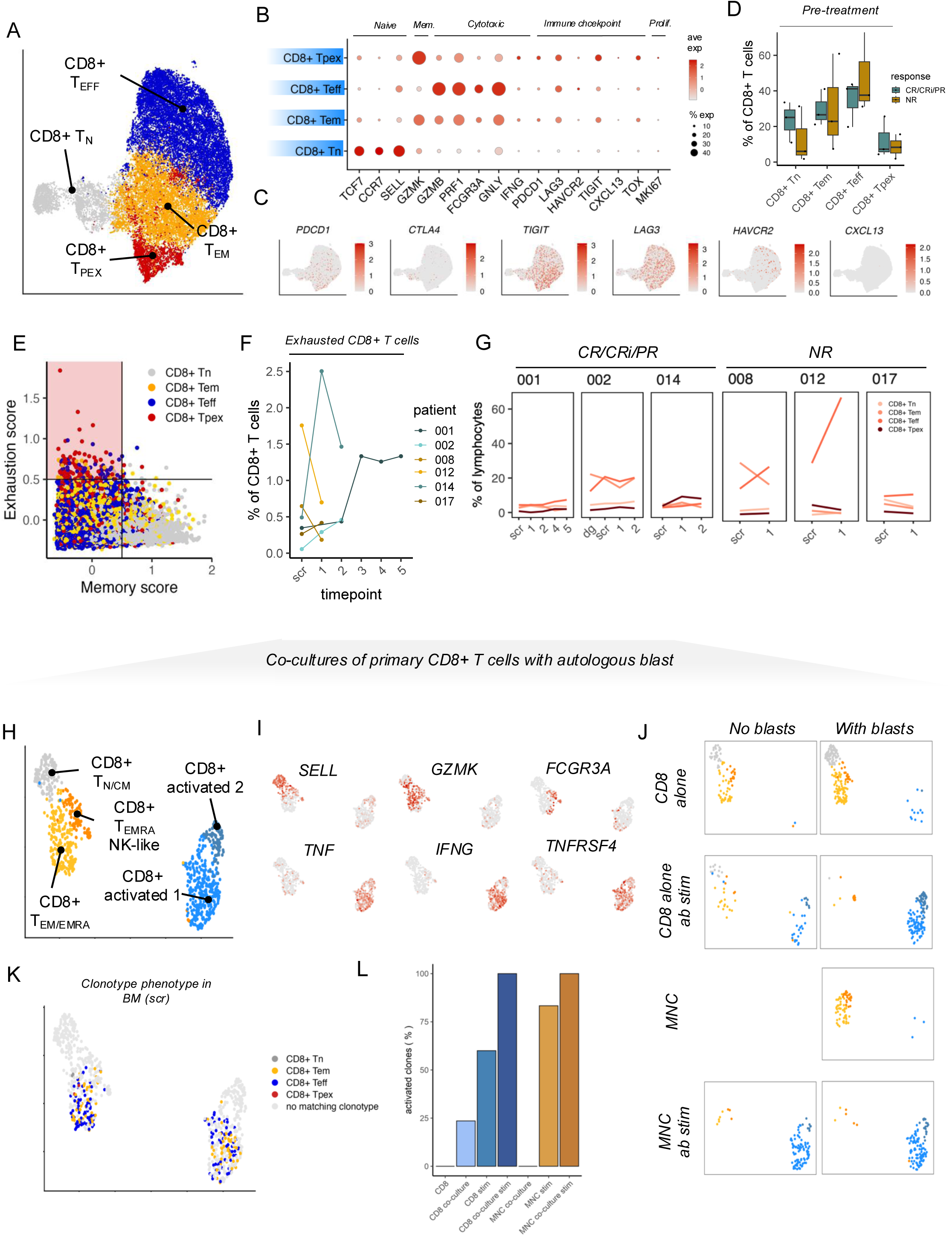
CD8+ T cell responses to anti-TIM3+HMA treatment. (A) UMAP of bone marrow CD8+ T cells from AML/MDS scRNA-seq profiles (*n*=7 patients, 20 samples) colored by manually annotated CD8+ T cell subtypes. (B) Average expression (color) and percentage expression (dot size) of canonical marker genes for CD8+ T cell subtype annotation. (C) Expression of selected checkpoint molecules in CD8+ T cells in projection from panel B. (D) Percentage of CD8+ T cell subtypes (Teff, Tn, Tem, Tpex) in pre-treatment bone marrow samples from responders (CR/CRi/PR) vs. non-responders (NR). (E) Relationship between exhaustion score and memory score for individual CD8+ T cells, colored by subtype. (F) Percentage of total CD8+ T cells within lymphocytes over treatment time points for individual patients. (G) Proportion of CD8+ T cell subtypes within total CD8+ T cells over treatment time points, comparing patient response groups (CR/CRi/PR vs. NR). (H) UMAP of CD8+ T cells from patient 001 CD8+ T cells when co-cultured with blast and MNCs with and without T cell stimulations. (I) Expression of selected phenotype markers in CD8+ T cells in projection from panel H. (J) Same UMAP as in panel H showing the distribution of cells across different co-culture conditions. (K) Same UMAP as in panel H highlighting the clones found both in co-culture and in the bone marrow data and the phenotype in bone marrow. (L) The proportion of CD8+ T cells belonging to activated clusters in different co-culture conditions.

We did not observe high gene expression levels of exhaustion markers in the pre-treatment BM aspirate samples within our cohort (**Fig. 4C**), consistent with findings from previous scRNA-seq analyses^4,17,39^. To ensure that our clustering approach did not underestimate the presence of exhausted T cells, we performed a clustering-independent analysis based on gene expression scoring to validate the number of exhausted cells in each sample^4^. This orthogonal analysis confirmed our findings, as we identified a median of 0.419% CD8+ T cells expressing canonical exhaustion molecules in pre-treatment samples (**Fig. 4E**). The proportion of exhausted cells was slightly higher in non-responders compared to responders (0.65% vs 0.35%), with the highest level observed in non-responder 012 (1.76%) (**Fig. 4F**). Returning to our cluster-based analysis, we did not observe a significant expansion of the CD8+ T cell pool in responders but noted such an expansion in two non-responders (**Fig. 4G**).

### CD8+ T cells are capable of recognizing autologous AML blasts

In functional co-culture assays, CD8+ T cells from patient 001 were able to respond to CD3+CD28+CD49d stimulation, demonstrating two activation states (expression of e.g. *TNF, IFNG,* and *TNFRSF4* encoding CD134/OX40R) upon co-culture, further confirming that at least not a majority of CD8+ T cells are exhausted (**Fig. 4H-L, Supplementary Fig. 10A-B**). Strikingly, when co-cultured with autologous blasts, CD8+ T cells transitioned more into activated states (**Fig. 4J**). As expected, the shift to activation state was more pronounced when stimulated with CD3+CD28+CD49d (**Fig. 4J**).

To pinpoint which phenotypes become activated following co-culture with blasts, we used TCRs as natural barcodes to map phenotypes from the BM to the co-culture data (**Fig. 4K**). Approximately 24% of CD8+ T cell clones (4 out of 17 detected clonotypes) were activated in the co-culture of MNCs and AML blast cells, even in the absence of CD3+CD28+CD49d antibody mediated T cell stimulation (**Fig. 4L**). Most clonotypes that became activated had a CD8+ T_EM_/T_EFF_ phenotype in the BM, and not e.g., NK-like T_EMRA_ or exhausted phenotype (**Supplementary Fig. 10C-D**).

### Anti-TIM3+HMA preferentially expands small CD8+ T cell clones

Next, we analyzed the TCR repertoires in more detail to identify which cells may be responding to anti-TIM3+HMA therapy. We measured TCR clonality with Gini index, where higher values denote more clonal populations. As expected, the baseline clonalities were higher in CD8+ than in CD4+ T cells (**Fig. 5A**). Responding patients had higher CD4+ T cell clonality than non-responders, best exemplified by patient 001 carrying the CD4+ T-LGLL clone. In contrast, the non-responders had a higher CD8+ T cell clonality at baseline (**Fig. 5A**), which is the opposite of what has been observed in anti-PD1 responders with metastatic melanoma^40^.

**Figure 5:**
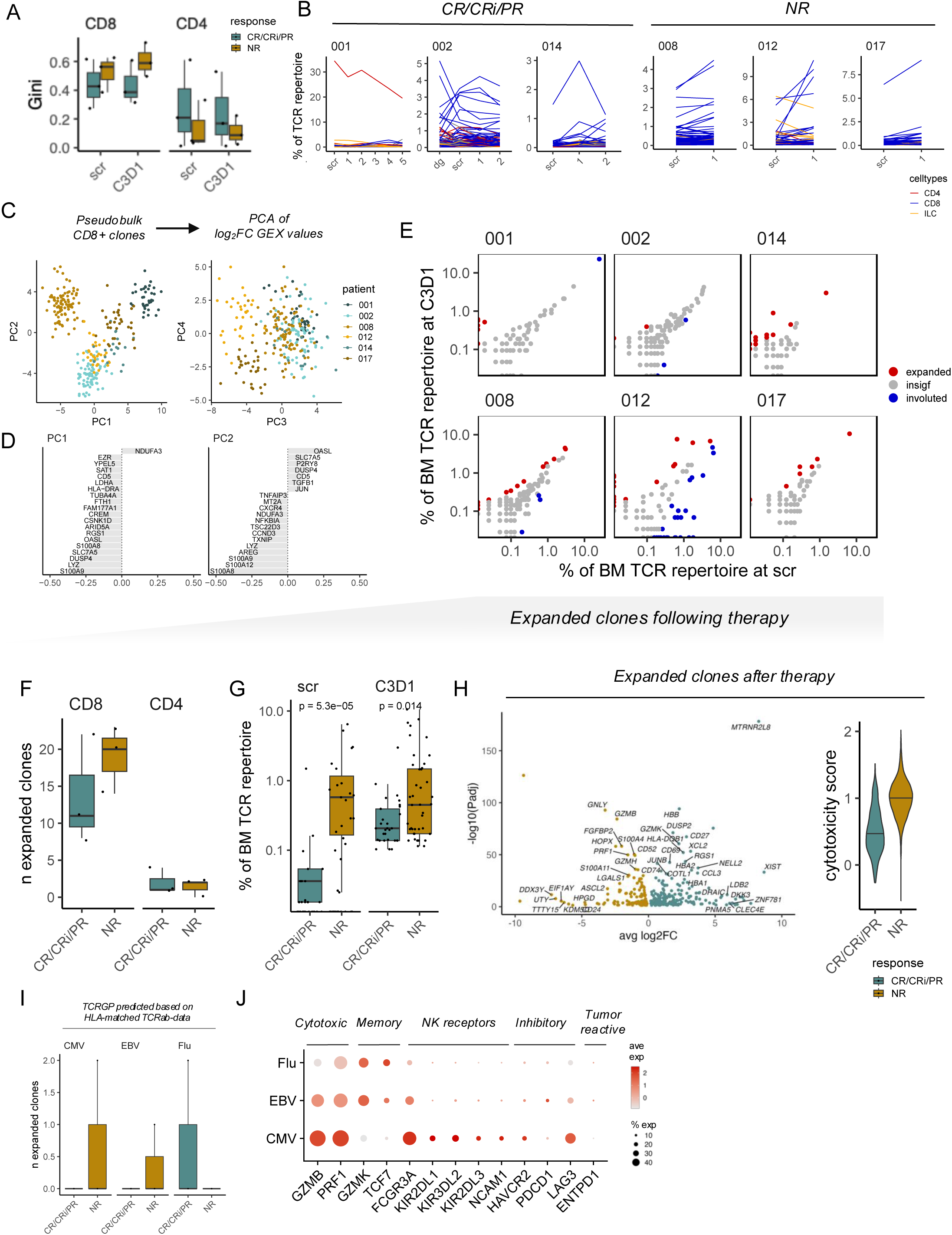
TCR repertoire dynamics and clonal expansion after anti-TIM3+HMA treatment. (A) TCR clonality (Gini index) for CD8+ and CD4+ T cells in bone marrow at baseline (C0D1) and C3D1, comparing responders (CR/CRi/PR) vs. non-responders (NR). (B) Proportion of major T cell subtypes (colors) within the TCR repertoire across treatment time points for individual patients. (C) Principal component analysis (PCA) of pseudobulk CD8+ T cell clones (left) and log2 fold change of gene expression values within these clones (right), colored by patient. (D) Top gene loadings for principal components 1, 2, and 3 from panel C. (E) Percentage of bone marrow TCR repertoire occupied by individual clones at C3D1 vs. screening (scr) for representative patients, highlighting expanded (red) and other clones. (F) Number of expanded CD8+ and CD4+ T cell clones at C0D1 and C3D1, comparing responders vs. non-responders. (G) Size (percentage of BM TCR repertoire) of expanded CD8+ and CD4+ T cell clones at screening and C3D1, comparing responders vs. non-responders. P-values from Mann-Whitney U-test. (H) Gene expression differences (DEGs) in CD8+ T cell clones expanded after therapy. (I) Number of expanded T cell clones with predicted specificity to CMV, EBV, or Influenza A based on HLA-matched TCRβ-seq data. (J) Average expression (color) and percentage expression (dot size) of genes related to cytotoxicity, memory, NK receptors, inhibitory molecules, and tumor reactivity in expanded T cell clones.

Although the overall clonalities remained similar following therapy, we found clonotypes that expanded following therapy in every patient (**Fig. 5B**). We wanted to identify putative tumor-reactive CD8+ T cells and first performed an unbiased analysis of clones following therapy. We performed pseudobulking of each clonotype identified in both pre- and post-therapy samples. The greatest variance in the data was explained by the individual patients from whom the clones originated (**Fig. 5C-D**), which hindered the identification of tumor-reactive clones based on transcriptomic data.

Therefore, we adopted an alternative approach by focusing on clones that significantly expanded after therapy, as these are likely enriched for tumor-specific targets. We identified a median of 11 statistically significantly expanded CD8+ T cell clonotypes in responding patients and 20 clones in non-responding patients (**Fig. 5E-F**, see **Methods**) and far less among CD4+ T cells. The most expanded clonotypes were identified in non-responding patients 008 and 012, while patient 014, with a slow response, also had multiple expanded clonotypes that kept expanding following C3D1 (**Fig. 5E**). In patient 001, we found four clones that expanded after anti-TIM3+HMA therapy also in the co-culture data, but low cell numbers hindered analysing these in detail (**Supplementary Fig. 10E-F**).

In the responding patients, most of the expanded CD8+ T cell clones were of low abundance in pre-treatment samples (mostly <0.1% of the TCR repertoire), unlike in the non-responding patients (*P*=5.3×10^−5^, Mann-Whitney U test, **Fig. 5G**). After treatment, the clones were still larger in non-responding patients (*P*=0.014, **Fig. 5H**). Unexpectedly, the expanded clones in non-responders after therapy exhibited a more cytotoxic phenotype than those in responders, characterized by higher expression of *GZMB/H*, *GNLY*, and NK cell receptors *FCGR3A*/CD16 and *KLRD1*, along with elevated cytotoxicity scores (**Fig. 5H**). This effect was especially noted in patient 012, who had the expansion of the largest clonotypes. This patient had particularly high expression of several immune checkpoint molecules (*HAVCR2/*TIM3*, PDCD1/*PD1*, CTLA4, LAG3, TIGIT*) in lymphocytes (**Supplementary Fig. 3B**) and experienced multiple infections during treatment, alongside a lack of clinical response (**Supplementary Data 1**).

T cells recognizing latent viral targets, such as those from herpes viruses, also express immune checkpoints like *HAVCR2*/TIM3 and may be modified by immune checkpoint inhibitors. Therefore, we predicted anti-viral TCRs with our machine learning method TCRGP^41^ with stringent criteria focusing on specificity rather than sensitivity (**Methods**, false-positive rate of 0%). Owing to HLA-restrictions, we could predict specificities for responding patients 002, 014, and non-responders 012, 017. Non-responder 012 had the highest number of anti-viral TCRs, especially targeting CMV, and three of its anti-CMV clones expanded following therapy (**Fig. 5I, Supplementary Fig. 11A-B)**. These anti-CMV T cells expressed cytotoxic genes (*GMZB, PRF1*), NK cell receptors (*FCGR3A* encoding CD16, different KIRs), and inhibitory molecules (*HAVCR2*, *PDCD1*, *LAG3*) (**Fig. 5J**).

### Myeloid cell phenotypes in anti-TIM3+HMA treatment

Finally, we focused on myeloid cells. We identified 8 different myeloid cell phenotypes from the scRNA+TCRαβ-seq data (**Fig. 6A**). The leukemic cells were identified based on the haplotype-aware copy number variation (CNV) analysis in patients that had those: i.e., 001 was marked by deletion 7 (del(7)), patient 009 with deletion 5q (del(5q)), and 017 with complex karyotype (del(5q), del(7q), marker chromosomes) (**Fig. 6A)**. We also attempted to identify malignant cells using somatic nuclear variants (SNVs), but, as in previous studies, we were unable to confidently resolve them due to biased coverage of short reads towards the 5’ prime end.

**Figure 6:**
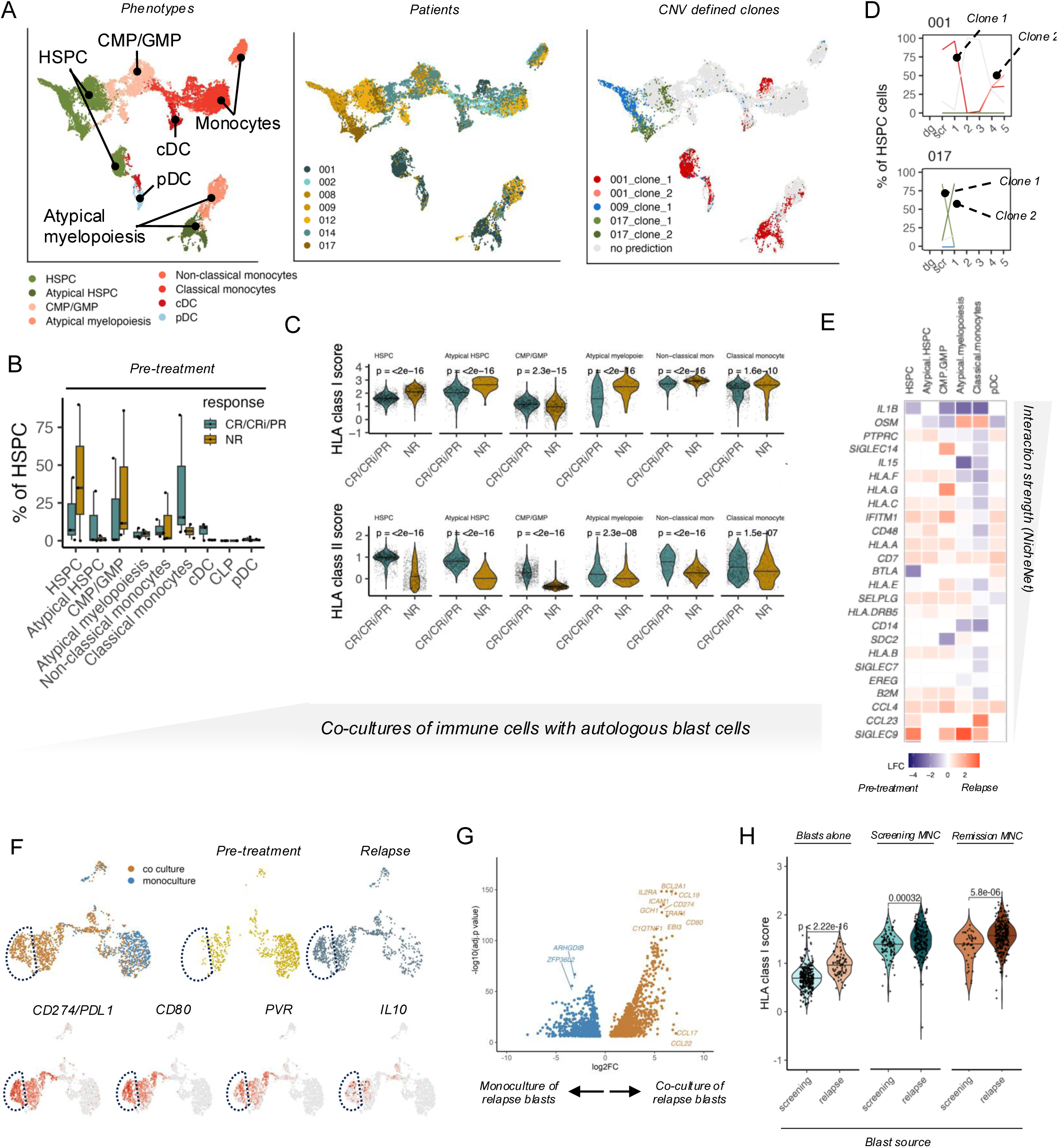
Myeloid cells responses to anti-TIM3+HMA treatment and immune escape mechanisms. (A) UMAP of bone marrow myeloid and hematopoietic stem/progenitor cells (HSPC) from AML/MDS scRNA-seq profiles, colored by cell phenotype (left), patient origin (middle), or CNV-defined malignant clones (right). (B) Percentage of myeloid/HSPC subtypes in pre-treatment bone marrow samples, comparing responders (CR/CRi/PR) vs. non-responders (NR). (C) HLA class I and HLA class II gene expression score distributions across myeloid/HSPC subtypes in pre-treatment samples, comparing responders vs. non-responders. (D) Differential gene expression (Log2 fold change) of selected genes in myeloid subsets comparing pre-treatment vs. relapse samples. (E) Differential gene expression (Log2 fold change) of significant ligand-receptor genes between myeloid cells and NK cells comparing pre-treatment versus relapse samples in patient 001. The values presented here are expression of ligands in myeloid cells. (F) Expression of selected genes *CD274*, CD80, PVR, and IL10 on myeloid cells (UMAP) comparing pre-treatment and relapse samples for an illustrative patient. (G) Differentially expressed genes in relapse blasts either in monoculture or in co-culture with patient MNCs. (H) Comparison of blast signature score, MHC class I/II expression (Screening MHC), and relapse MHC expression distributions in malignant cells.

In the scRNA+TCRαβ-seq data, responding patients had a higher proportion of more differentiated myeloid cells, like classical monocytes, than non-responders at baseline (**Fig. 6B**). This was also noted in flow cytometry analysis, where the number of CD16+ monocytes remained higher in responders’ PB before and after treatment (**Supplementary Fig. 11C**).

Downregulation of HLA molecules may confer resistance to T cell mediated therapies but create vulnerability to NK cell mediated therapies^7^. Prior to treatment, responders had lower HLA class I scores than non-responders in their myeloid cells in scRNA+TCRαβ-seq cohort (**Fig. 6C**). Interestingly, responders had higher HLA class II scores than non-responders in their myeloid cells, irrespective of the analyzed subtype (**Fig. 6C**).

### Patient cases show signs of immune evasion

Patient 001 achieved CR after the first cycle and minimal residual disease (MRD) negativity before cycle 6 day 1 (C6D1) as their best response (**Supplementary Note 1**). Based on the CNV analysis, the malignant clones at screening showed varying phenotypes from HSPC to monocytes and dendritic cells (**Fig. 6A**). After stable MRD negativity of two years, BM sample from C19D23 showed MRD positivity (fluorescence in situ hybridization (FISH) for del(7): 2/1000 interphases), but C20D6 was MRD negative again. However, C24D2 showed a morphological relapse with MRD positivity (FISH 2.1%). In the scRNAseq data, we could observe an emerging new clone with an additional CNV gain in chromosome 19 that phenotypically resembled mature myeloid cells (**Supplementary Fig. 12A, Fig. 6D**).

We studied the interactions between NK cells and the clone dominant at relapse and clone dominant at screening with NicheNet^42^, where the emerging clone had an upregulation of class I HLA-molecules (*HLA-A, HLA-B, HLA-F*) and *B2M* (**Fig. 6E**) that likely confers resistance to NK cells^7^. One of the genes that had the highest predicted interaction between blast cells and NK and CD4+ T-LGLL cells, *IL1B*, was lost in the new clone (**Fig. 6E**). In previous studies, *IL1B* has been identified as one the main mediators of anti-TIM3 response^43^.

To further investigate the blast cells from the relapse time point, we compared the malignant CD34+ blasts at screening and relapse co-cultured with MNCs. Interestingly, relapse blasts upregulated cell-cycle genes, like *CCND1*, and immune evasion genes like *IL10*, and immune checkpoint ligands such as *CD274* (encoding PD-L1) and *PVR* (ligand for *TIGIT)* in the co-culture conditions (**Fig. 6F-G**). Also, relapse blasts exhibited the highest levels of HLA class I expression in co-cultures (**Fig. 6H**).

Patient 017, with a strong adaptive NK cell response, had a *TP53^mt^*complex karyotype MDS, that previously failed azacytidine therapy and was enrolled to receive sabatolimab as monotherapy. With CNV profiling at single-cell level, we could reliably follow the malignant cells. During the treatment, the patient’s adaptive NK cells showed promising response by expansion and a decrease in the cells in clone 1 (**Fig. 6D**). During the treatment course, we could observe an enrichment of a new malignant clone with additional del(2q), which had a HSPC phenotype (clone 2, **Fig. 6D**, **Supplementary Fig. 13A-B**). The DEG analysis between the previously dominating clone 1 and the new dominant clone 2 suggested upregulation of intrinsic apoptotic resistance and cell cycle arrest (e.g., *CDKN1A, CDKN2A, CDKNC2,* and *BCL2*), potentially offering an escape from adaptive NK cell immunity (**Supplementary Fig. 13C-E**).

## Discussion

Here, we report the first translational study of cancer patients treated with anti-TIM3 in combination with decitabine in conjunction to a phase Ib trial (NCT03066648)^16^. Our study highlights unique aspects of how TIM3 blockade combined with HMA may modulate immune responses in myeloid malignancies, a process likely driven by the broad expression of *HAVCR2*/TIM3 across immune cell types.

One of our key findings is the impact of TIM3 blockade on NK cells, particularly on mature CD56^dim^ and adaptive NK cell subsets. We have previously shown that myeloid malignancies are among the most sensitive to NK cell mediated killing within hematological cancers^7^. Moreover, prior studies have revealed that TIM3 blockade enhances IFN-γ secretion and activation of NK cells, suggesting that this may be a key mechanism underlying the efficacy of anti-TIM3+HMA therapy. Other novel immune checkpoint inhibitor therapies, such as anti-LAG3+anti-PD1 combination, also demonstrate reinvigoration of adaptive NK cells during treatment^44^.

Interestingly, responders to anti-TIM3+HMA therapy exhibited a greater oligoclonal expansion of cytotoxic CD4+ T cells compared to non-responders. TIM3 was originally identified as a marker of cytotoxic CD4+ cells, underscoring its potential role in modulating this subset. Supporting this, a murine model with conditional TIM3 knockout showed a CD4+ phenotype similar to our observations^23^. The most striking example of oligoclonal cytotoxic CD4+ T cells was observed in the exceptional responder who had a pre-existing diagnosis of CD4+ T-LGLL, a condition that typically follows an indolent course with autoimmune manifestations such as neutropenia, as seen in this patient. T-LGLL has been speculated to represent an extreme immune response to a persistent antigen; however, to our knowledge, no specific target antigens have been identified in T-LGLL patients. In this study, we confirmed that the CD4+ T-LGLL clone harbored a TCR reactive to autologous leukemic cells, providing insight into the potential origin of T-LGLL clones. Our previous work demonstrated that T-LGLL induces immune dysregulation beyond the malignant clone, including increased clonality, elevated IFN-γ expression, and enhanced antigen presentation and class II expression in myeloid cells^9^, all of which were evident in this patient. These findings support the intriguing possibility that mild autoimmunity against self-antigens may enhance response to immune checkpoint blockade, similar to the association between vitiligo and favorable outcome in melanoma immunotherapy^45^.

Anti-TIM3+HMA therapy appeared to preferentially expand small, rather than large, CD8+ T cell clones in responders. This suggests that anti-TIM3 may primarily modulate the tumor microenvironment, such as antigen-presenting cells, rather than directly reversing CD8+ T cell exhaustion. This finding contrasts with previous murine studies where TIM3 blockade enhanced CD8+ T cell activation^23^, although TIM3 knockout did not lead to similar improvements in CD8+ T cell mediated anti-tumor immunity^52^. Notably, large CD8+ T cell clones were more common in non-responders and were associated with immune-related adverse effects, consistent with observations from anti-PD1+HMA therapy in AML^3^. One possible explanation for the limited CD8+ T cell mediated response to anti-TIM3+HMA is the scarcity of endogenous CD8+ T cells targeting AML/MDS presenting epitopes, as reflected by the low frequency (<1%) of exhausted CD8+ T cells and lack of response to anti-PD1/anti-CTLA4. Importantly, this does not diminish the potential of adoptive cell therapies based on CD8+ T cells or NK cells retargeted with AML-specific markers, such as WT1, CD123, or recurrent neoantigens. On the contrary, it may further support the rationale for outsourcing the T cell response through such strategies.

Finally, in the leukemic cells, we observed widespread HLA dysregulation, a well-established mechanism of immune escape following allogenic transplantation. We also identified evidence of clonal evolution associated with acquired resistance, including the emergence of additional CNVs in patient 001 and enrichment of an apoptosis-resistant clone in patient 017. The impact of TIM3 on myeloid cells may represent an additional mechanism of action for TIM3-based therapies. We look forward to the results of post-HSCT treatment strategies administrated prior to morphologic relapse, which aim to enhance the graft-versus-leukemia effect.

The primary limitations of our study stem from the phase I design of the clinical trial, which involved a limited number of patients. We addressed this constraint through systematic sample collection and in-depth functional profiling of the most informative cases, particularly the exceptional responders. As the study did not include a cohort receiving HMA monotherapy, we could not definitively attribute observed effects solely to anti-TIM3. However, previous studies have extensively characterized HMA-treated patients using flow cytometry and scRNA-seq. Functional analyses were limited to the exceptional responder, patient 001, due to sample availability. Nevertheless, based on correlative data, we believe this patient provided the most insight into the treatment’s putative mechanism of action.

Our findings support a role for anti-TIM3+HMA therapy in enhancing anti-leukemia immunity, particularly through the activation of NK cells and CD4+ T cells in AML/MDS. These results suggest that this treatment approach is an interesting option in selected patient subgroups, such as those with clonal CD4+ T cells and/or adaptive NK cell populations.

## Materials and methods

### Study design

We performed an in-depth immunomonitoring in the phase Ib trial (NCT03066648)^16^ of decitabine in combination with anti-TIM3 (sabatolimab, MBG453). This study was approved by the Helsinki University Hospital ethics committee, and the principles of Helsinki Declaration were followed. All patients had given their written informed consent before sample collection. No compensation was provided for the study participants. No statistical methods were used to predetermine sample size. The experiments were not randomized, and the investigators were not blinded to allocation during experiments and outcome assessment.

### AML/MDS patients

We enrolled 13 patients with newly diagnosed or relapsed or refractory AML unfit to intensive chemotherapy and 1 patient with MDS to the phase I trial. The treatment started with 1 priming cycle of decitabine monotherapy (20 mg/m2 on days 1-7) followed by treatment using a combination of decitabine and sabatolimab (3-10 mg/m2 on day 1, (**Figure 1A**). The newly diagnosed patients with AML were unfit for intensive chemotherapy or declined from it, and the relapsed and/or refractory patients with AML were treated with different regimens. Prior to therapy, the leukocyte numbers were reduced with hydroxyurea (*n*=5). Adverse effects that were deemed immunological were treated with corticosteroids such as methylprednisolone, prednisone, or hydrocortisone (*n*=6). The clinical characteristics are detailed in **Supplementary Data 1.**

We routinely collected PB from screening, following priming cycle of decitabine (C1D8), and then on first day of every cycle and BM aspirate samples from screening and on first day of every third cycle and end of treatment (EOT).

### Single-cell RNA+TCRαβ sequencing

We subjected 20 samples from 7 patients to scRNA+TCRαβ-seq. As we wanted to focus on the effects on the lymphoid populations, we enriched for lymphocytes by profiling a mixture of 90% lymphocytes and 10% myeloid cells, including the putative blast cells (representative gating in **Supplementary Fig. 1A**)

Single-cells were partitioned using a Chromium Controller (10x Genomics) and scRNA+TCRαβ-seq and TCRαβ-libraries were prepared using Chromium Single Cell 5’ Library & Gel Bead Kit (10x Genomics), as per manufacturer’s instructions (CG000086 Rev D). The gene expression libraries were sequenced with Illumina NovaSeq and the TCR-enriched libraries with Illumina HiSeq2500.

Data processing was performed using 10x Genomics Cell Ranger v3.0.1 pipeline, where “cellranger mkfastq” was used to produce FASTQ (raw data) files, “cellranger count” to perform alignment, filtering and UMI counting for the 5’ gene expression data and “cellranger vdj” to perform V(D)J sequence assembly and paired clonotype calling for the V(D)J data. mkfastq was run using the Illumina bcl2fastq v2.2.0 and alignment was done against human genome GRCh38.

Cells with high number of mitochondrial transcripts (>15% of all UMI counts), high amount of ribosomal transcripts (>50%), low or high number of detected genes (<100 or >4500), or low or high read depth (<500 or >30,000) were excluded from the analyses. Potential batch-effects were accounted for with scVI-tools^46^ (v 0.5.0) with default parameters where each sample was treated as a batch as per best practices^47^. The batch-corrected latent embeddings from scVI were then used for UMAP-dimensionality reduction^48^ and graph-based clustering implemented in Seurat^49^ (v 4.3.0) with default parameters.

To get the most optimal representation of cell phenotypes, we used a reclustering scheme by identifying broader cell lineages (CD8+, CD4+, Tunconv, NK, B, myeloid and HSPC) by overclustering the data and then subjected these more detailed cell lineages to reclustering (**Supplementary Fig. 1B**). First, the cells were separated into myeloid cells and lymphoid cells. From the lymphoid cells, B cells were extracted, and the following T/NK cells were subjected to additional overclustering and identification of canonical T cells and innate lymphocyte cells based on expression of TCR, TCR-constant gene expression, and careful annotation of *CD3E/D/G* expression. CD4+ and CD8+ T cells were delineated by expression of *CD8A/B*, cytotoxic markers (e.g., *GZMA/B/M/H*, *PRF1*), and clonal lineage analyses (i.e., if most cells from a TCR clonotype had been identified as CD8+, the rest of the clone was identified as CD8+). The resulting broader cell type lineages (CD8+, CD4+, Tunconv, NK, B, myeloid, and HSPC) were then subjected to final clustering, where the latent embeddings were used for the default graph-based clustering with the Leiden method implemented in Seurat (v 4.3.0), with default parameters and a resolution parameter 1.0. Clusters were annotated by analyzing canonical markers, DEGs, relationship to other clusters, different signature scores, and automated reference-based Celltypist^50^ (v 1.2.0) and SingleR^51^ (v 2.20) predictions.

DEGs were identified with Student’s *t*-test and subsequent pathway enrichment analyses were performed with hypergeometric testing on GO, HALLMARK, Reactome, and TF categories from ClusterProfiler (v. 2.1.4). Gene signature scores were calculated with AddModuleScore-function with default parameters^52^. The genes used to calculate the cytotoxicity score included *IFNG*, *GZMA*, *GZMB, GZMM, GZMH, PRF1,* and *GNLY*; HLA class I score included *HLA-A, HLA-B,* and *HLA-C*; HLA class II score included *HLA-DMA*, *HLA-DMB*, *HLA-DOA*, *HLA-DOB*, *HLA-DPA1*, *HLA-DPB1*, *HLA-DQA1*, *HLA-DQA2*, *HLA-DQB1*, *HLA-DQB1-AS1*, *HLA-DQB2*, *HLA-DRA*, *HLA-DRB1*, and *HLA-DRB5;* exhaustion score: *CTLA4*, *ENTPD1*, *HAVCR2*, *LAG3*, *PDCD1*, *TIGIT*, *TOX,* and *CXCL13;* memory score: *CCR7*, *CD28*, *IL7R*, *SELL*, and *TCF7*. Intercellular communications were studied with NicheNet^42^ (v. 2.2.0) with default parameters on an individual patient level. Pseudotime analyses were studied with Slingshot^34^ (v. 3.2.2) on UMAP reductions from latent embeddings with default parameters.

For patients with clinically detected copy number variations (CNVs; 001, 009, 017), we performed haplotype-aware somatic CNV analysis using Numbat (v 1.3.0) on each patient separately with default parameters. T and NK cells were designated as the CNV-negative reference group, and the myeloid cells were subjected to CNV detection. Various reference groups were tested, including T and NK cells, T cells alone, and the Human Cell Atlas reference provided with Numbat.

TCR clones were defined as having the same identical, nucleotide level TCRαβ-sequence from the CellRanger output with custom scripts. Clonality was assessed with the Gini inequality metric from ineq R-package (v 0.2-13). For supervised detection of epitope-specific clonotypes, TCRGP^41,54^ (v. 1.0.0) for CMV, EBV, and Influenza A specific data were trained using 10-fold cross-validation with default parameters with 2000 iterations and a learning rate of 0.005. Only predictions with a false-positive rate of 0% were retained, and only TCRs with only one predicted target were considered as significant.

### Immunophenotyping with flow cytometry and analysis

Frozen BM and PB samples were immunophenotyped with flow cytometry using 5 panels to obtain different immune cell subpopulations and TIM3 (TIM3-BB515 (565569), CD4-PE-Cy7 (560649), GammaDelta-APC (555718)); immune-checkpoint receptors (PD1-FITC (557860), LAG3-PE (12-2239-42, eBiosciences), ICOS-PE-Cy7 (25-9948-41, eBiosciences), CTLA4-APC (560938)); NK-cell and activation markers (NKG2C-AF488 (FAB138G, R&D Systems), CD161-PE (556081), NKG2D-PE-Cy7 (562365), NKG2A-APC (FAB1059A, R&D Systems) and DNAM-BB515 (565152), CD57-PE (560844), NKp46-PE-Cy7 (562101), NKp30-Alexa647 (558408)) and markers for migration and cell memory status (CXCR3-AF488 (561730), CCR7-PE (FAB197P, R&D Systems), CD45RO-PE-Cy7 (560608), CXCR4-APC (560936)). In addition, each panel included markers for main immune cell subpopulations (CD45-APC-H7 (560178), CD3-PerCP-Cy5.5 (332771), CD56-BV421 (562751), CD8-BV510 (563919)). All antibodies were purchased from BD Biosciences unless otherwise indicated. Samples were acquired with FACS Verse (BD), and the data were analyzed with FlowJo (FlowJo 10.8, FlowJo).

### Generation of T-LGLL Jurkat cells

Most abundant clonotype TCR alpha and beta chains from patient 001 CD4+ T-LGLL were constructed from scRNA+TCR sequencing data and ordered as synthetic DNA from IDT (Integrated DNA Technologies), cloned into a lentivirus vector using Gibson Assembly (New England Biolabs) to be transduced into Jurkat 6.1 T cells, which is a Jurkat model that is engineered to be negative for TCR alpha and beta chains and to express CD8.

HEK293FT cells were seeded in RPMI-1640 supplemented with 10% FBS and 2 mM L-glutamine and incubated at 37°C with 5% CO₂ for 24 hours. For transfection complex preparation, Opti-MEM was mixed with Lipofectamine 2000 and incubated for 5 minutes at room temperature (20°C). Separately, TCR plasmid, psPAX2, and pMD2.G were diluted in Opti-MEM. The DNA mixture was combined with the Lipofectamine solution and incubated for 25 minutes at room temperature. Subsequently, DNA–Lipofectamine complex was added to the HEK293FT cells, followed by incubation at 37°C with 5% CO₂ for 48–72 hours.

Viral supernatant was harvested, centrifuged at 300 × g for 5 minutes, filtered through a 0.45 µm membrane, and applied to Jurkat 6.1 reporter T cells in the presence of 8 µg/mL polybrene. Transduction was enhanced by spinoculation (800 × g, 2 hours). Following centrifugation, the viral medium was aspirated and cells were resuspended in fresh RPMI-1640 with 10% FBS and 2 mM L-glutamine. Jurkat cells were incubated at 37°C with 5% CO₂ for 72 hours. TCR transgene expression was confirmed by antibody staining for TCR and CD3 followed by their flow cytometry analysis. A parallel strategy was used to generate a control Jurkat cells devoid of exogenous TCR expression (mock Jurkat cells).

### Co-culture experiments of CD34+ blast, mononuclear cells, and T-LGLL Jurkat cells

For functional experiments, patient 001 BMMCs from pre-treatment screening of AML and relapse were thawed and sorted for CD34+ blasts using an aCD34 bead kit (Miltenyi Biotech, Cat. 130-046-702). Patient BMMCs from AML remission were also thawed but not sorted due to negligible blast percentage. The CD34+ blast and CD34-MNC populations were rested overnight at 3M/mL in cRPMI with 5%CO2 at 37°C. T-LGLL Jurkat reporter T cells were likewise thawed and rested in cRPMI at 1M/mL. Next, the CD34+ blasts and CD34-MNC were co-cultured with T-LGLL Jurkat reporter T cells or each other using an E:T ratio of 1:1 overnight. For T cell stimulation, antibodies were added to select wells: anti-CD3 (5.55μg/ml; clone UCHT1, BD, Cat. 555335), anti-CD28 (1μg/ml), and anti-CD49d (1 μg/ml). Different culture conditions were pooled with TotalSeq C hashtag antibodies (Biorad) and processed for scRNA+TCRαβ-seq.

### Co-culture data analysis

To classify cells per condition, sequenced cells underwent demultiplexing of hashtag oligonucleotide (HTO) counts. Centered log-ratio-normalized HTO UMI counts were used with the “HTODemux”-function in Seurat (v. 5.2.1) with a uniquely chosen positive quantile for each sample, ranging from 0.99 to 0.9999. For primary co-cultures cells with high number of mitochondrial transcripts (>15% of all UMI counts) or ribosomal transcripts (>50%), cells with less than 150 genes or over than 10.000 genes expressed, and cells with low or high read depth (<4000 or > 80.000) were excluded from the analyses. Remaining doublets were identified using “hybrid score” from scds (v.3.1.0) with a hybrid score > 1.6. For engineered T cell co-cultures, cells with less than 1000 genes or over than 7500 genes expressed, and cells with low or high read depth (<2000 or >40 000) were excluded from the analyses.

Batch corrected latent embeddings with scVI-tools^46^ (v 0.5.0) was used to combine data from all conditions. In the model, default parameters were used. Each condition was treated as a batch and cell cycle as a covariate. The latent embeddings were then used for UMAP-dimensionality reduction in Seurat with default parameters.

Different immune cell lineages were delineated with manually curated markers, with TCR information and unsupervised clustering, as described before. After delineating each immune cell lineage was re-clustered to better define cell subtypes. Only cells classified as singlets and cell types corresponding to the experimental design (e.g., only myeloid lineage cells from CD34+ sorted samples) were retained for further analyses.

### Statistical testing

*P*-values were calculated with nonparametric tests, including Mann-Whitney U test (two groups), Kruskal-Wallis test (more than two groups), and Fisher’s exact test, where the alternative hypotheses are reported. *P*-values were corrected with Benjamini-Hochberg adjustment. All calculations were done with R (4.3.0) or Python (3.9.13).

## Data Availability

The scRNA+TCRαβ-seq data can be received from Zenodo 10.5281/zenodo.16754663 with a restricted access due to GDPR regulations, and data can be accessed by placing a request via Zenodo. The custom scripts to reproduce the key findings can be found in https://github.com/janihuuh/tim3_manu.

https://zenodo.org/records/16754663

## Acknowledgments

We are deeply grateful to all patients who participated in the study and generously contributed samples.

This project was funded by the grants from Novartis Pharma, Sigrid Juselius Foundation, Signe and Ane Gyllenberg Foundation, Jane and Aatos Erkko Foundation, Research Council of Finland, Helsinki Institute for Life Science, and Cancer Foundation Finland. J.H. was supported by Blood Disease Research Foundation, Biomedicum Helsinki Foundation, Finnish Medical Foundation, K. Albin Johansson Foundation, Kaute Foundation, Suomalais-Norjalainen Lääketieteen Säätiö, and Emil Aaltonen Foundation.

We acknowledge IT Center for Science Ltd for data storage and computational resources and the computational resources provided by the Aalto Science-IT project. The flow cytometry analysis was performed at the HiLife Flow Cytometry Unit, University of Helsinki. scRNA+TCRαβ-seq was performed at FIMM Single-Cell Analytics unit supported by HiLIFE and Biocenter Finland.

## Author Contributions Statement

JH: Conceptualization, methodology, software, formal analysis, investigation, data curation, writing – original draft, visualization, project administration.

SF: Investigation, methodology, formal analysis, data curation, writing – original draft, visualization.

BF: Investigation, methodology, formal analysis, data curation, writing – original draft, visualization.

JS: Investigation, methodology, formal analysis, data curation.

OB: Conceptualization, methodology, software, investigation, formal analysis, data curation.

SL: Methodology.

AK: Conceptualization, investigation, data curation.

OD: Investigation.

MK: Investigation.

MI: Conceptualization, investigation, data curation.

HLäht: Investigation.

TK: Investigation.

JK: Investigation.

JL: Methodology

PS: Methodology, resources

MKo: Resources.

MP: Resources.

HLähd: Investigation, supervision.

MR: Resources.

CS-P: Resources.

KP: Conceptualization, resources, project administration.

KaP: Conceptualization, investigation, formal analysis, data curation, project administration.

SM: Conceptualization, investigation, resources, writing – original draft, supervision, project administration, funding acquisition.

All authors read and approved the final manuscript.

## References

1. Penter, L. et al. Molecular and cellular features of CTLA-4 blockade for relapsed myeloid malignancies after transplantation. Blood 137, 3212–3217 (2021).

2. Abbas, H. A. et al. Single cell T cell landscape and T cell receptor repertoire profiling of AML in context of PD-1 blockade therapy. Nat Commun 12, 6071 (2021).

3. Goswami, M. et al. Pembrolizumab and decitabine for refractory or relapsed acute myeloid leukemia. J Immunother Cancer 10, (2022).

4. Penter, L. et al. Mechanisms of response and resistance to combined decitabine and ipilimumab for advanced myeloid disease. Blood (2023).

5. Papaemmanuil, E. et al. Genomic Classification and Prognosis in Acute Myeloid Leukemia. N Engl J Med 374, 2209–2221 (2016).

6. Schumacher, T. N. & Schreiber, R. D. Neoantigens in cancer immunotherapy. Science 348, 69–74 (2015).

7. Dufva, O. et al. Single-cell functional genomics reveals determinants of sensitivity and resistance to natural killer cells in blood cancers. Immunity 56, 2816–2835.e13 (2023).

8. Lundgren, S. et al. Single-cell analysis of aplastic anemia reveals a convergence of NK and NK-like CD8+ T cells with a disease-associated TCR signature. Sci Transl Med 17, eadl6758 (2025).

9. Huuhtanen, J. et al. Single-cell characterization of leukemic and non-leukemic immune repertoires in CD8+ T-cell large granular lymphocytic leukemia. Nat. Commun. 13, 1981 (2022).

10. Wolf, Y., Anderson, A. C. & Kuchroo, V. K. TIM3 comes of age as an inhibitory receptor. Nat Rev Immunol 20, 173–185 (2020).

11. Kikushige, Y. et al. A TIM-3/Gal-9 Autocrine Stimulatory Loop Drives Self-Renewal of Human Myeloid Leukemia Stem Cells and Leukemic Progression. Cell Stem Cell 17, 341–352 (2015).

12. Haubner, S. et al. Coexpression profile of leukemic stem cell markers for combinatorial targeted therapy in AML. Leukemia 33, 64–74 (2019).

13. Wei, A. H. et al. Sabatolimab (MBG453) Dose Selection and Dose-Response Analysis in Myelodysplastic Syndrome (MDS)/Acute Myeloid Leukemia (AML): Population Pharmacokinetics (PK) Modeling and Evaluation of Clinical Efficacy/Safety By Dose. Blood 136, 40–42 (2020).

14. Zeidan, A. M. et al. Sabatolimab plus hypomethylating agents in previously untreated patients with higher-risk myelodysplastic syndromes (STIMULUS-MDS1): a randomised, double-blind, placebo-controlled, phase 2 trial. Lancet Haematol 11, e38–e50 (2024).

15. Brunner, A. M. et al. Phase Ib study of sabatolimab (MBG453), a novel immunotherapy targeting TIM-3 antibody, in combination with decitabine or azacitidine in high- or very high-risk myelodysplastic syndromes. Am J Hematol 99, E32–E36 (2024).

16. Schwartz, S., et al. Characterization of sabatolimab, a novel immunotherapy with immuno-myeloid activity directed against TIM-3 receptor. Immunother Adv 2, ltac019 (2022).

17. Dufva, O. et al. Immunogenomic Landscape of Hematological Malignancies. Cancer Cell 38, 424–428 (2020).

18. Petti, A. A. et al. A general approach for detecting expressed mutations in AML cells using single cell RNA-sequencing. Nat Commun 10, 3660 (2019).

19. van Galen, P. et al. Single-Cell RNA-Seq Reveals AML Hierarchies Relevant to Disease Progression and Immunity. Cell 176, 1265–1281.e24 (2019).

20. Penter, L. et al. Integrative genotyping of cancer and immune phenotypes by long-read sequencing. Nat Commun 15, 32 (2024).

21. Savola, P., Bhattacharya, D. & Huuhtanen, J. The spectrum of somatic mutations in large granular lymphocyte leukemia, rheumatoid arthritis, and Felty’s syndrome. Semin Hematol (2022).

22. Dixon, K. O., Lahore, G. F. & Kuchroo, V. K. Beyond T cell exhaustion: TIM-3 regulation of myeloid cells. Sci Immunol 9, eadf2223 (2024).

23. Talvard-Balland, N. et al. Oncogene induced TIM-3 ligand expression dictates susceptibility to anti-TIM-3 therapy in mice. J Clin Invest (2024).

24. Mehtonen, J. et al. Single cell characterization of B-lymphoid differentiation and leukemic cell states during chemotherapy in ETV6-RUNX1-positive pediatric leukemia identifies drug-targetable transcription factor activities. Genome Med 12, 99 (2020).

25. Witkowski, M. T. et al. Extensive Remodeling of the Immune Microenvironment in B Cell Acute Lymphoblastic Leukemia. Cancer Cell 37, 867–882.e12 (2020).

26. Caron, M. et al. Single-cell analysis of childhood leukemia reveals a link between developmental states and ribosomal protein expression as a source of intra-individual heterogeneity. Sci Rep 10, 8079 (2020).

27. Zavidij, O. et al. Single-cell RNA sequencing reveals compromised immune microenvironment in precursor stages of multiple myeloma. Nat Cancer 1, 493–506 (2020).

28. Oetjen, K. A., et al. Human bone marrow assessment by single-cell RNA sequencing, mass cytometry, and flow cytometry. JCI Insight 3, (2018).

29. Sade-Feldman, M. et al. Defining T Cell States Associated with Response to Checkpoint Immunotherapy in Melanoma. Cell 175, 998–1013.e20 (2018).

30. Tang, F. et al. A pan-cancer single-cell panorama of human natural killer cells. Cell 186, 4235–4251.e20 (2023).

31. Huuhtanen, J. et al. Single-cell analysis of immune recognition in chronic myeloid leukemia patients following tyrosine kinase inhibitor discontinuation. Leukemia 38, 109–125 (2024).

32. Huuhtanen, J. et al. IFN-α with dasatinib broadens the immune repertoire in patients with chronic-phase chronic myeloid leukemia. J. Clin. Invest. 132, (2022).

33. Yang, C. et al. Heterogeneity of human bone marrow and blood natural killer cells defined by single-cell transcriptome. Nat Commun 10, 3931 (2019).

34. Street, K. et al. Slingshot: cell lineage and pseudotime inference for single-cell transcriptomics. BMC Genomics 19, 477 (2018).

35. Gumá, M. et al. Imprint of human cytomegalovirus infection on the NK cell receptor repertoire. Blood 104, 3664–3671 (2004).

36. Holmes, T. D., et al. The transcription factor Bcl11b promotes both canonical and adaptive NK cell differentiation. Sci Immunol 6, (2021).

37. Duàn, H. et al. Genomic Subtypes of AML Define Sensitivity to NK Cell Cytotoxicity. Blood 144, 329 (2024).

38. Schoggins, J. W. & Rice, C. M. Interferon-stimulated genes and their antiviral effector functions. Curr. Opin. Virol. 1, 519–525 (2011).

39. Mazziotta, F. et al. A phase I/II trial of WT1-specific TCR gene therapy for patients with acute myeloid leukemia and active disease post-allogeneic hematopoietic cell transplantation: skewing towards NK-like phenotype impairs T cell function and persistence. Nat Commun 16, 5214 (2025).

40. Tumeh, P. C. et al. PD-1 blockade induces responses by inhibiting adaptive immune resistance. Nature 515, 568–571 (2014).

41. Jokinen, E., Huuhtanen, J., Mustjoki, S., Heinonen, M. & Lähdesmäki, H. Predicting recognition between T cell receptors and epitopes with TCRGP. PLoS Comput. Biol. 17, e1008814 (2021).

42. Browaeys, R., Saelens, W. & Saeys, Y. NicheNet: modeling intercellular communication by linking ligands to target genes. Nat Methods 17, 159–162 (2020).

43. Dixon, K. O. et al. TIM-3 restrains anti-tumour immunity by regulating inflammasome activation. Nature 595, 101–106 (2021).

44. Huuhtanen, J. et al. Single-cell characterization of anti–LAG-3 and anti–PD-1 combination treatment in patients with melanoma. J. Clin. Invest. 133, (2023).

45. Tumor infiltrating lymphocytes (TIL) therapy in metastatic melanoma: boosting of neoantigen-specific T cell reactivity and long-term follow-up - PubMed. https://pubmed.ncbi.nlm.nih.gov/32753545/.

46. Lopez, R., Regier, J., Cole, M. B., Jordan, M. I. & Yosef, N. Deep generative modeling for single-cell transcriptomics. Nat Methods 15, 1053–1058 (2018).

47. Heumos, L. et al. Best practices for single-cell analysis across modalities. Nat Rev Genet 1–23 (2023).

48. McInnes, L., Healy, J. & Melville, J. UMAP: Uniform Manifold Approximation and Projection for Dimension Reduction. ArXiv StatML (2018).

49. Hao, Y. et al. Integrated analysis of multimodal single-cell data. Cell 184, 3573–3587.e29 (2021).

50. Domínguez Conde, C., et al. Cross-tissue immune cell analysis reveals tissue-specific features in humans. Science 376, eabl5197 (2022).

51. Aran, D. et al. Reference-based analysis of lung single-cell sequencing reveals a transitional profibrotic macrophage. Nat Immunol 20, 163–172 (2019).

52. Tirosh, I. et al. Dissecting the multicellular ecosystem of metastatic melanoma by single-cell RNA-seq. Science 352, 189–196 (2016).

53. Gao, T. et al. Haplotype-aware analysis of somatic copy number variations from single-cell transcriptomes. Nat Biotechnol 41, 417–426 (2023).

54. Huuhtanen, J. et al. Evolution and modulation of antigen-specific T cell responses in melanoma patients. Nat. Commun. 13, 5988 (2022).

