## Supplementary Material for "TIM3 blockade with hypomethylating therapy restores NK and cytotoxic CD4+ T cell activity in patients with AML or MDS"

Supplementary Figure 1

A

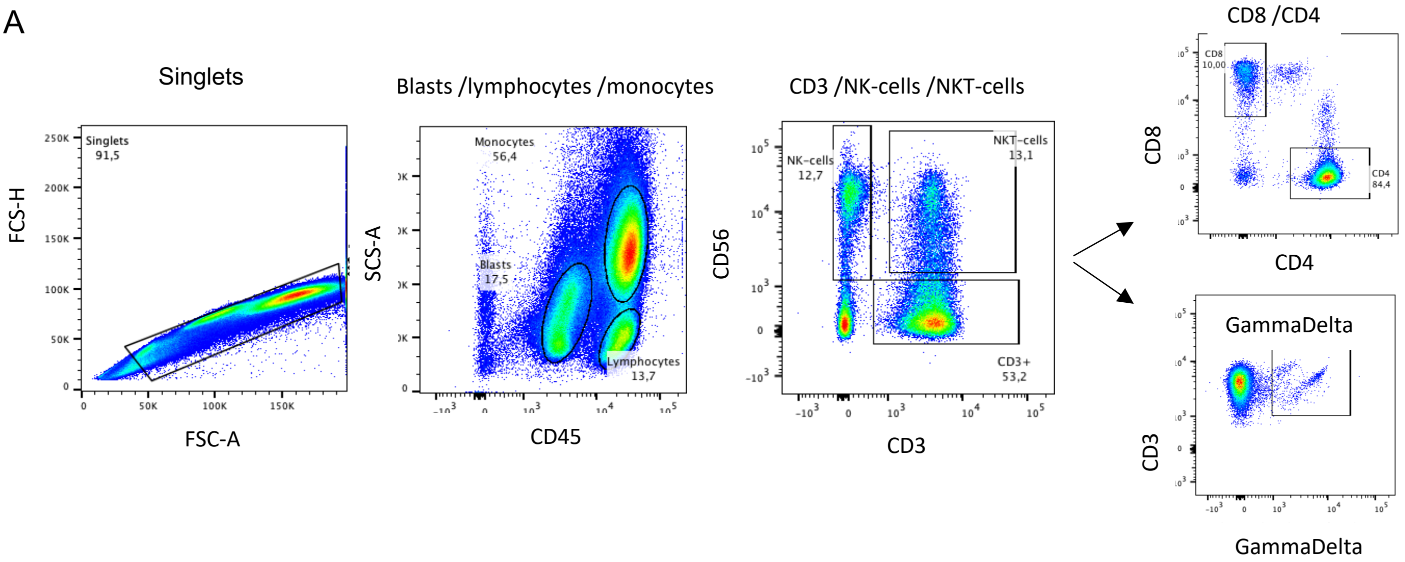


B

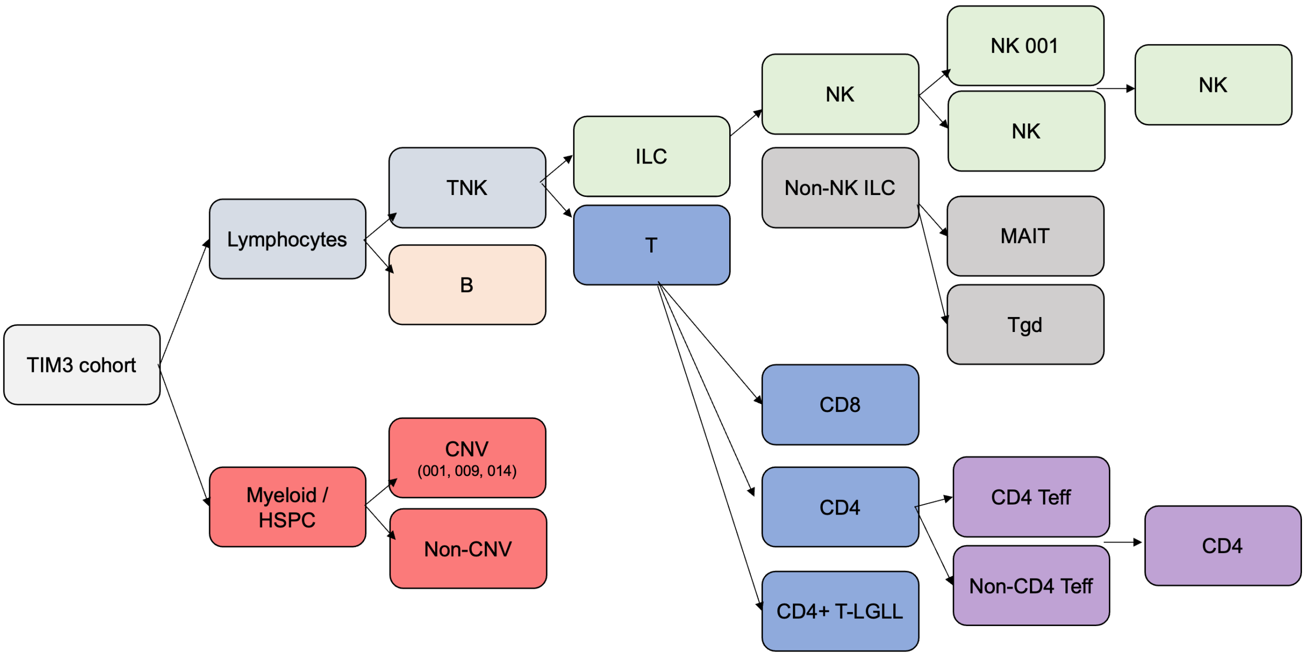

**Supplementary Figure 1: Study plan**

**A)** Example of flow cytometry gating strategy**.**
**B)** Flow chart of clustering strategies to identify the optimal number of cell types from scRNA-seq profiles.

Supplementary Figure 2

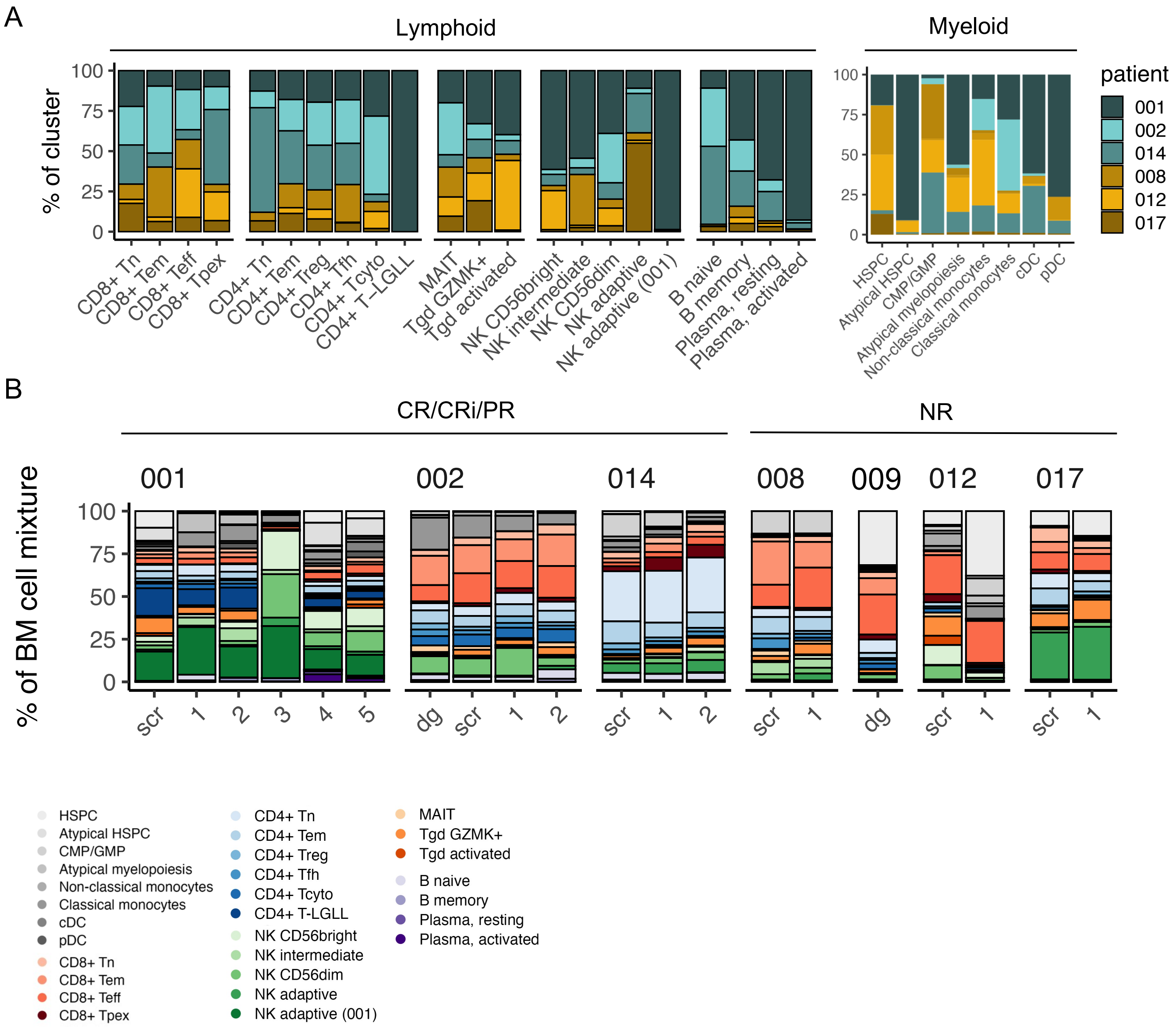

**Supplementary Figure 2: Single-cell RNA-seq cell type annotation**

**A)** Proportion of cells coming from different patients across cell types in scRNA-seq data.
**B)** Proportion of cells in different patients across time points in scRNA-seq data.

Supplementary Figure 3

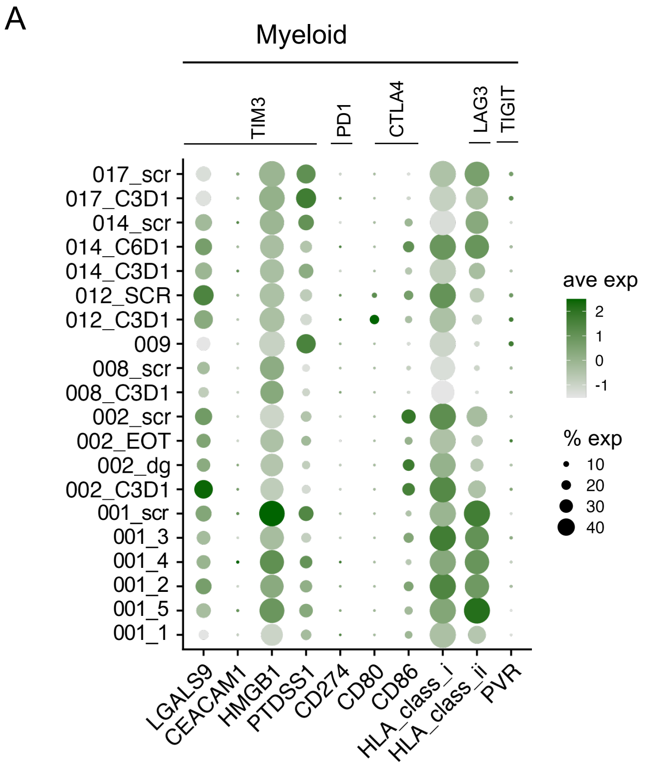


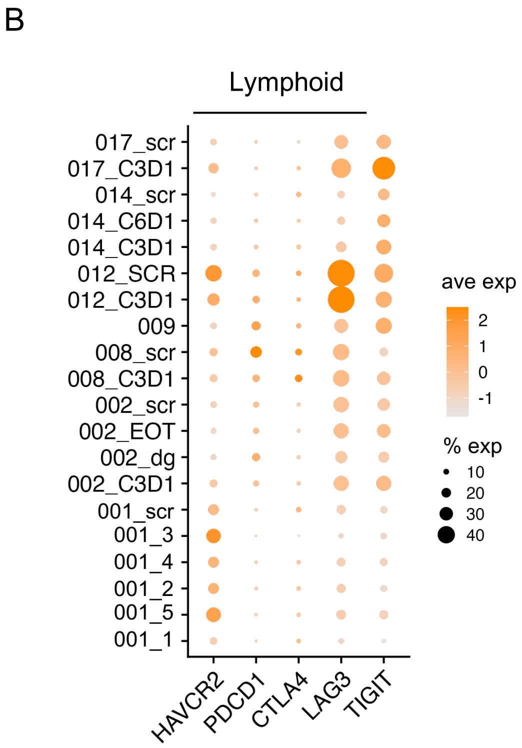

**Supplementary Figure 3: Immune checkpoint ligand and receptor expression**

**A-B**) Expression of key immune checkpoint ligands in myeloid cells **(A)** and receptors in lymphoid cells **(B)** in patient samples across time points in scRNA-seq data, where the size of the circle corresponds to the percentage of cells expressing the marker and the color to its average expression.

Supplementary Figure 4

A

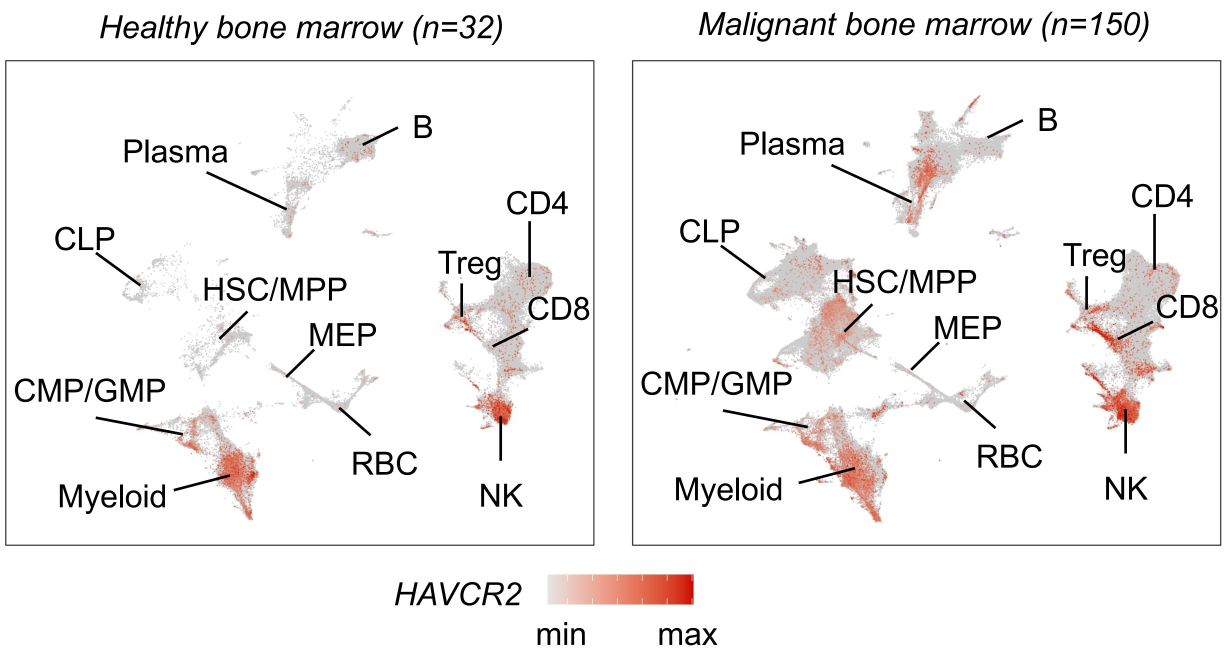


B

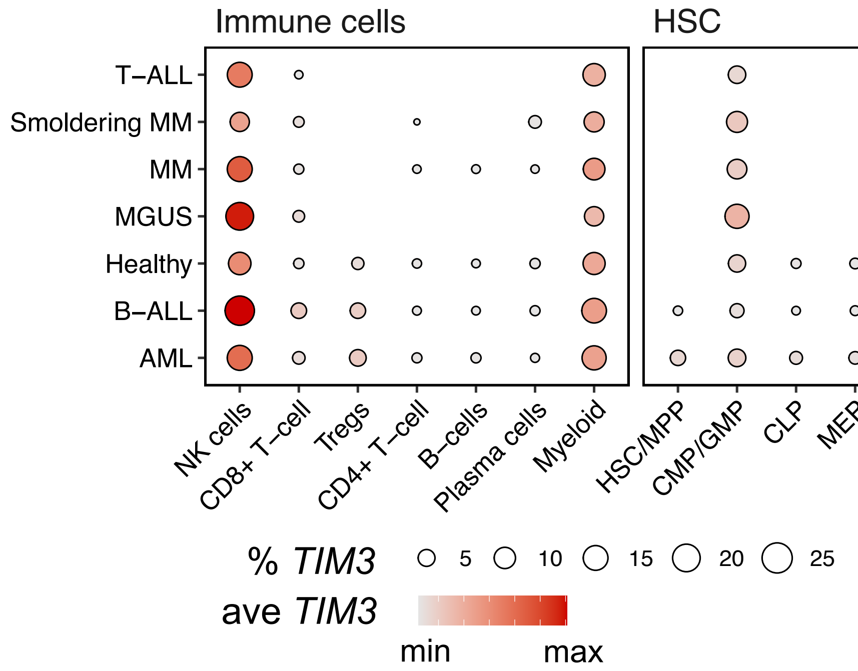

**Supplementary Figure 4: TIM3 expression**

**A)** UMAP showing the expression of *HAVCR2*/TIM3 in different cell types across healthy controls and 9 different hematological malignancies in scRNA-seq data.
**B)** Expression of *HAVCR2/*TIM3, where the size of the circle corresponds to the percentage of cells expressing the marker and the color to its average expression.

Supplementary Figure 5

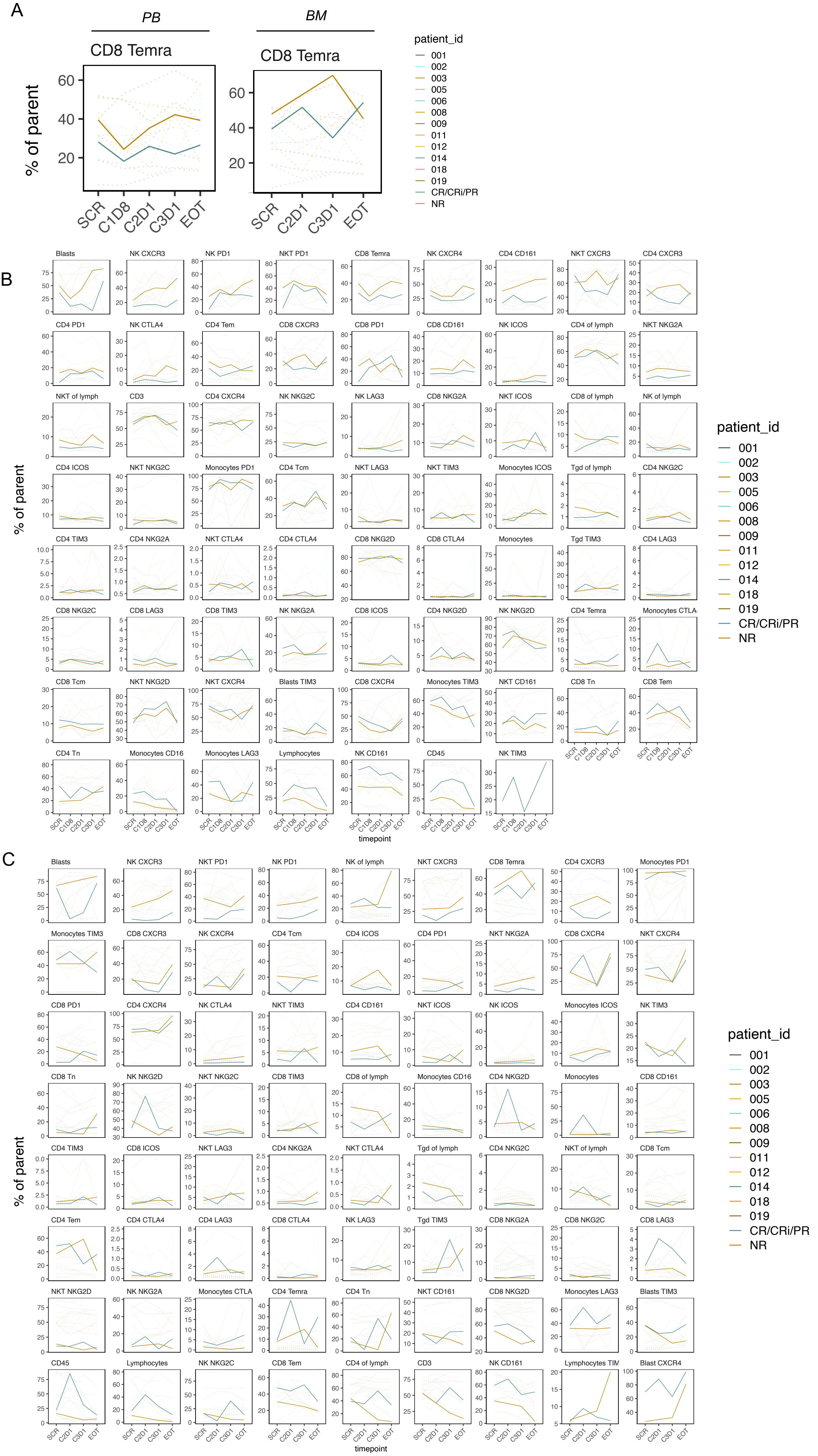

**Supplementary Figure 5: Flow cytometry data**

**A)** Proportion of CD8 $T_{EMRA}$ cells in peripheral blood and bone marrow as measured with flow cytometry.
**B-C)** Proportion of cells out of the parent population in flow cytometry samples from peripheral blood (B) and bone marrow (C). The dashed lines represent individuals and the highlighted line represents the average.

Supplementary Figure 6

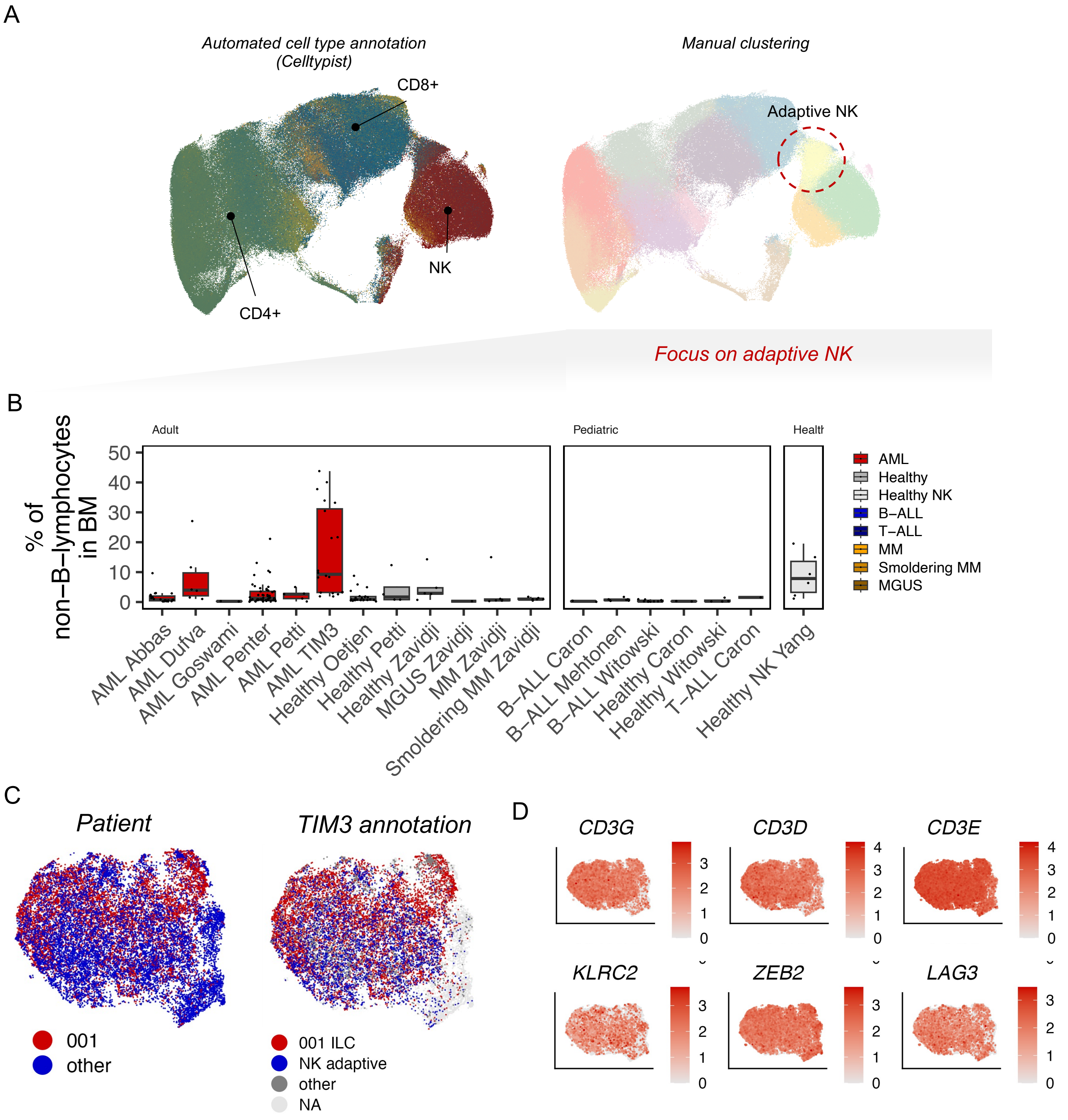

**Supplementary Figure 6: Definition of NK adaptive (001) cells**

**A)** UMAP of the T/NK from the pan-hematological malignancy cohort (*n*=182), where cells are colored either by an automated reference-based method Celltypist (left) or by clustering (right), where the putative adaptive NK cells are highlighted.
**B)** Proportion of putative adaptive NK cells from non-B-lymphocytes.
**C)** UMAP of the putative adaptive NK cells, where patient 001 is highlighted (left) or the cell type annotation used in Figure 1A (right).
**D)** Expression of canonical markers used to define adaptive NK cells.

Supplementary Figure 7

A

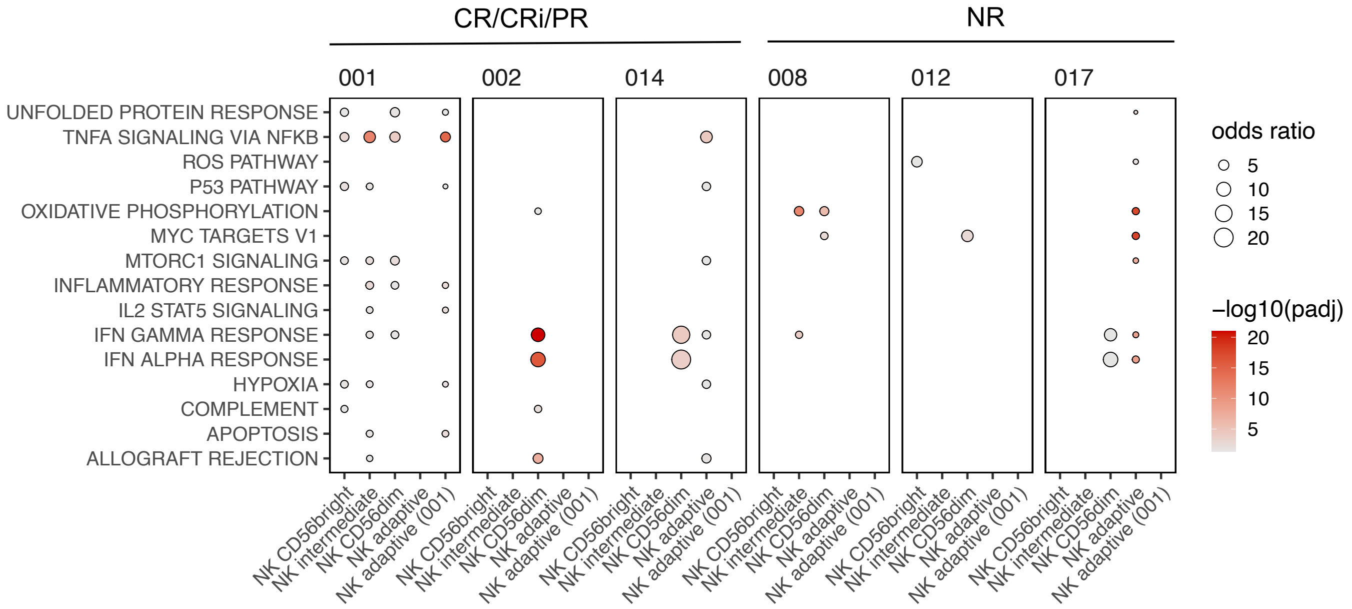

**Supplementary Figure 7: Effect of anti-TIM3+HMA on NK cell transcriptome**

**A)** Upregulation of different pathways following anti-TIM3+HMA treatment, where the size of the circle corresponds to the odds ratio of DE genes found in a category and the color to the findings adjusted *P*-value.

Supplementary Figure 8

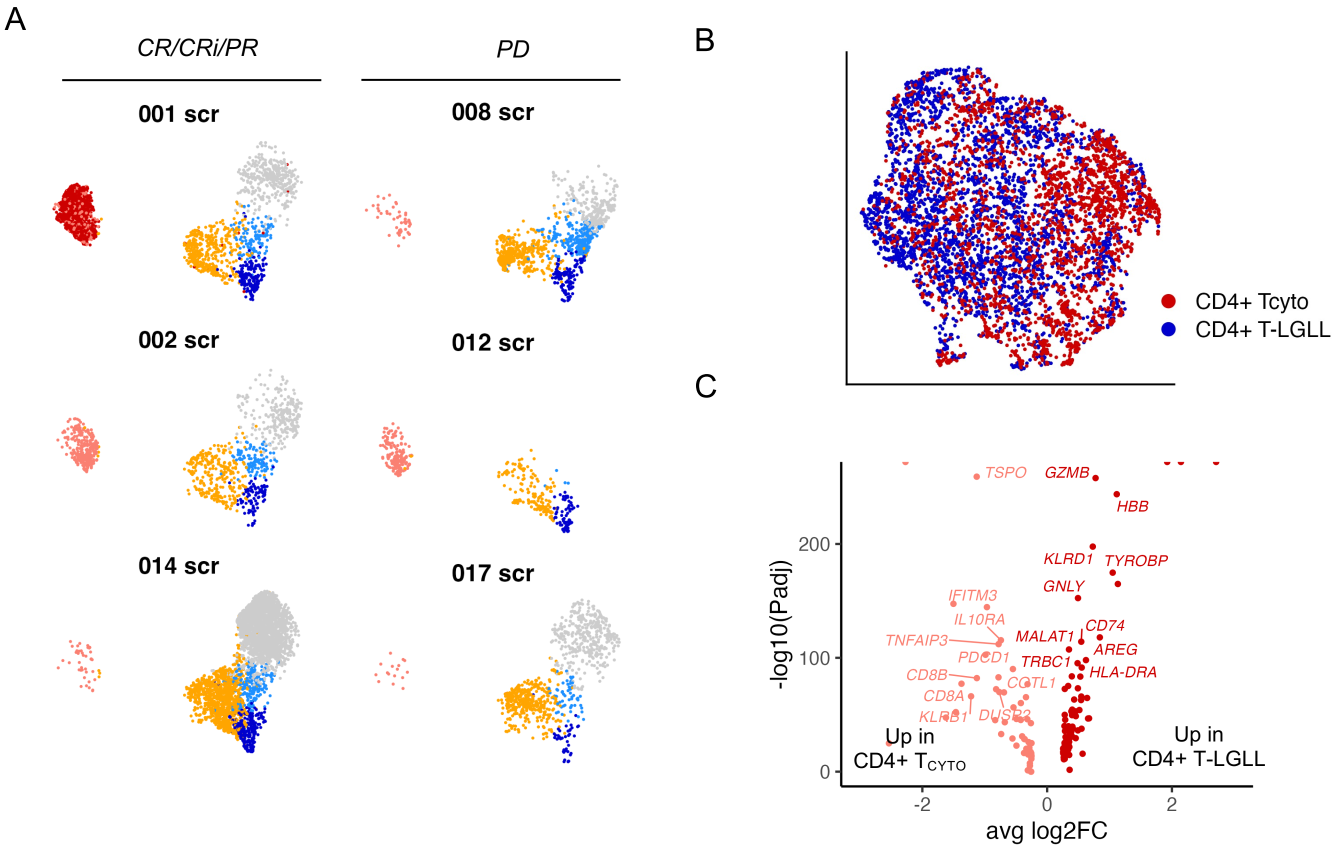

**Supplementary Figure 8: CD4+ T-LGLL cells**

**A)** UMAP of CD4+ T cells in different patients in pre-treatment samples.
**B)** UMAP of CD4+ T-LGLL cells and cytotoxic CD4+ T cells.
**C)** Differentially expressed genes between CD4+ T-LCLL and cytotoxic CD4+ T cells.

Supplementary Figure 9

A

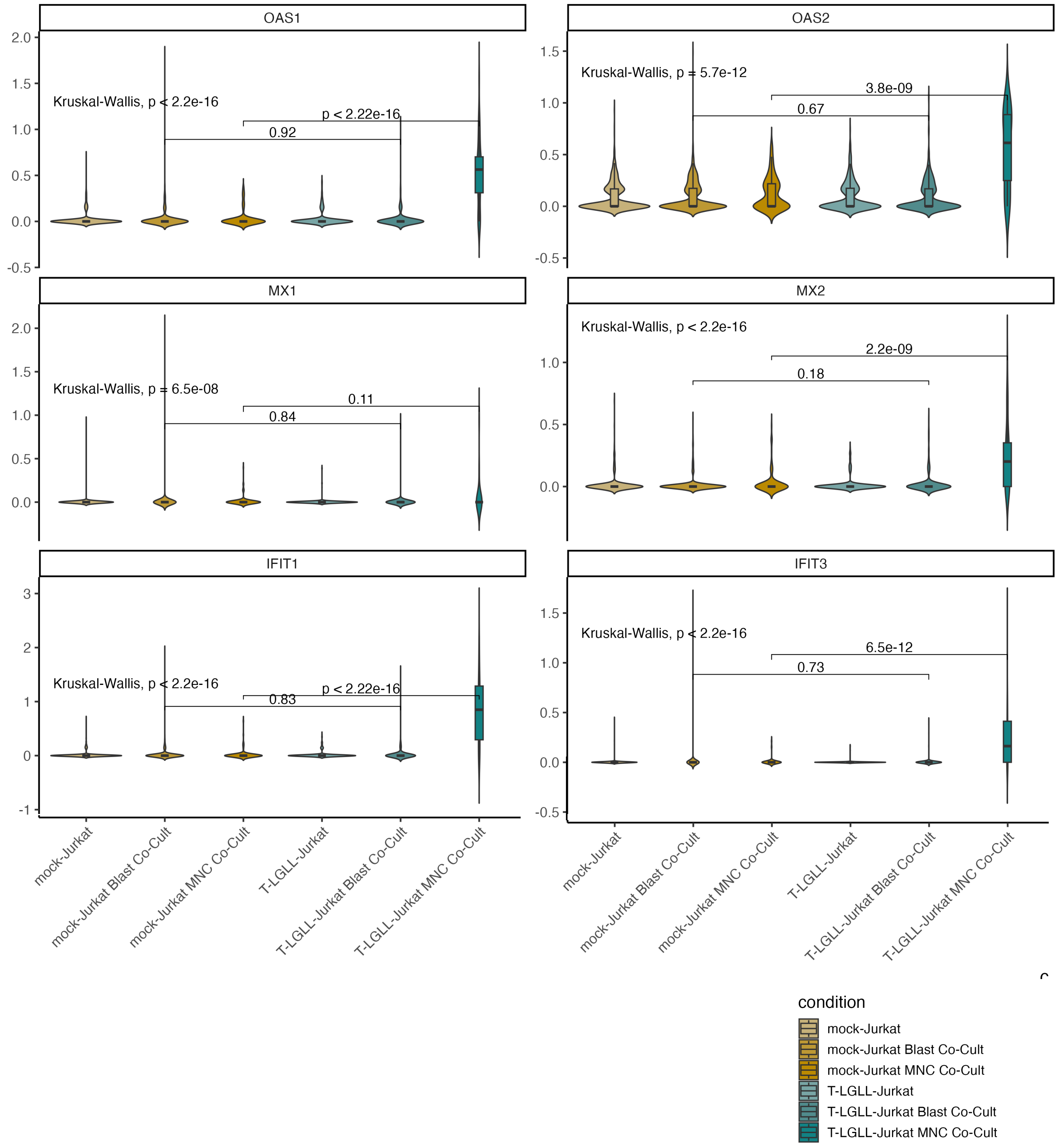

**Supplementary Figure 9: T-LGLL Jurkat co-culture experiments**

**A)** Expression of selected genes in T-LGLL Jurkat and mock-Jurkat co-cultures in different conditions, including monocultures and co-culture with autologous blasts and autologous mononuclear cells (MNCs). The experiments were performed without stimulating antibodies.

Supplementary Figure 10

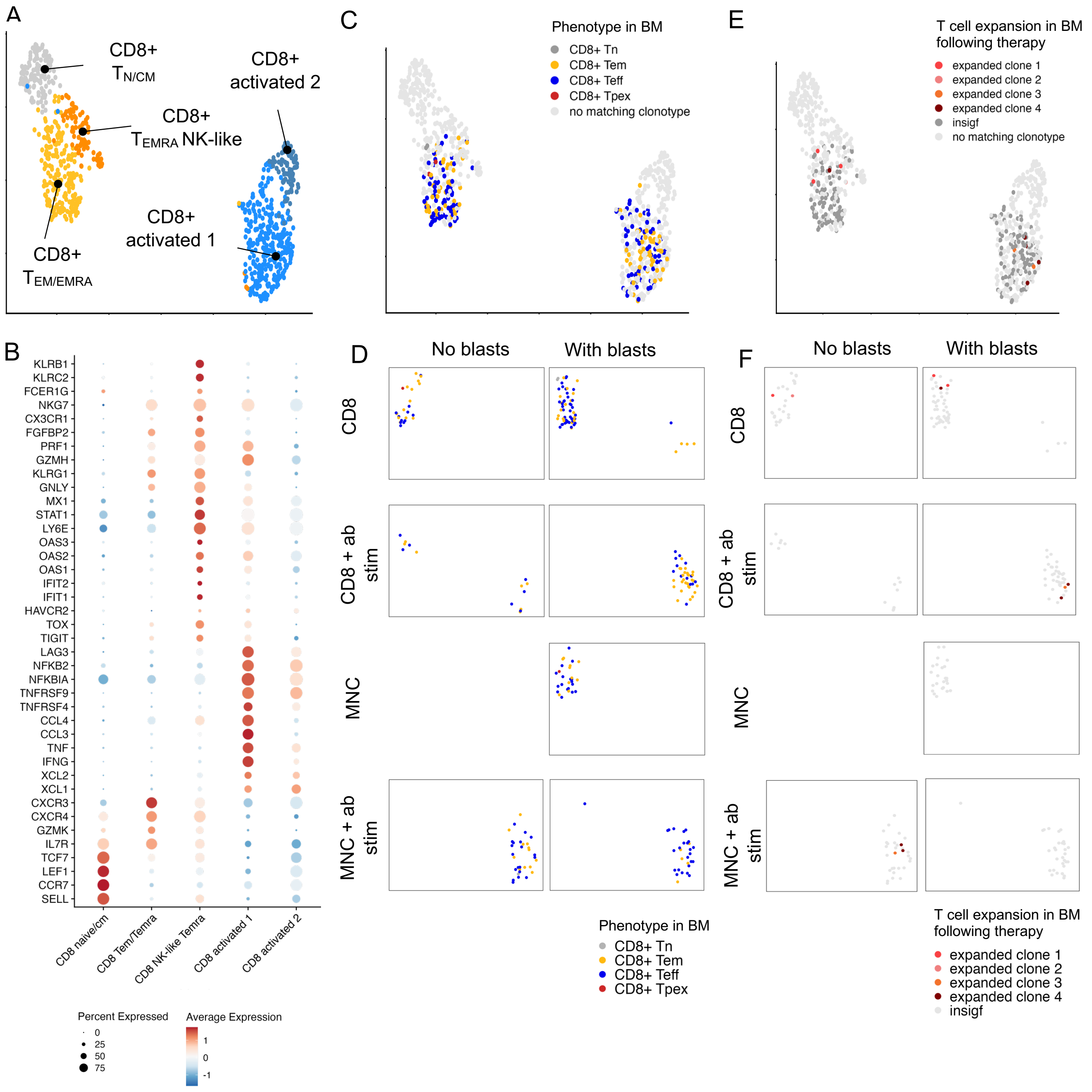

**Supplementary Figure 10: CD8+ T cells functional co-cultures from patient 001**

**A)** UMAP of CD8+ T cells from patient 001 from co-cultures in different conditions.
**B**) Expression of selected genes used for annotating CD8+ T cell clusters.
**C)** Same UMAP as in panel A highlighting the TCR clones found both in the co-culture in vitro scRNA+TCRαβ-seq data and the patient data from bone marrow scRNA+TCRαβ-seq data. The coloring highlights the dominating phenotype found in the bone marrow scRNA+TCRαβ-seq data.
**D)** Same UMAP as in panel C highlighting the different co-culture conditions. The coloring highlights the dominating phenotype found in the bone marrow scRNA+TCRαβ-seq data.
**E)** Same UMAP as in panel A highlighting the TCR clones found both in the co-culture in vitro scRNA+TCRαβ-seq data patient data from bone marrow and their expansion status following therapy. The coloring highlights whether the CD8+ T cell clone expanded significantly following therapy.
**F)** Same UMAP as in panel E highlighting the different co-culture conditions. The coloring highlights whether the CD8+ T cell clone expanded significantly following therapy.

Supplementary Figure 11

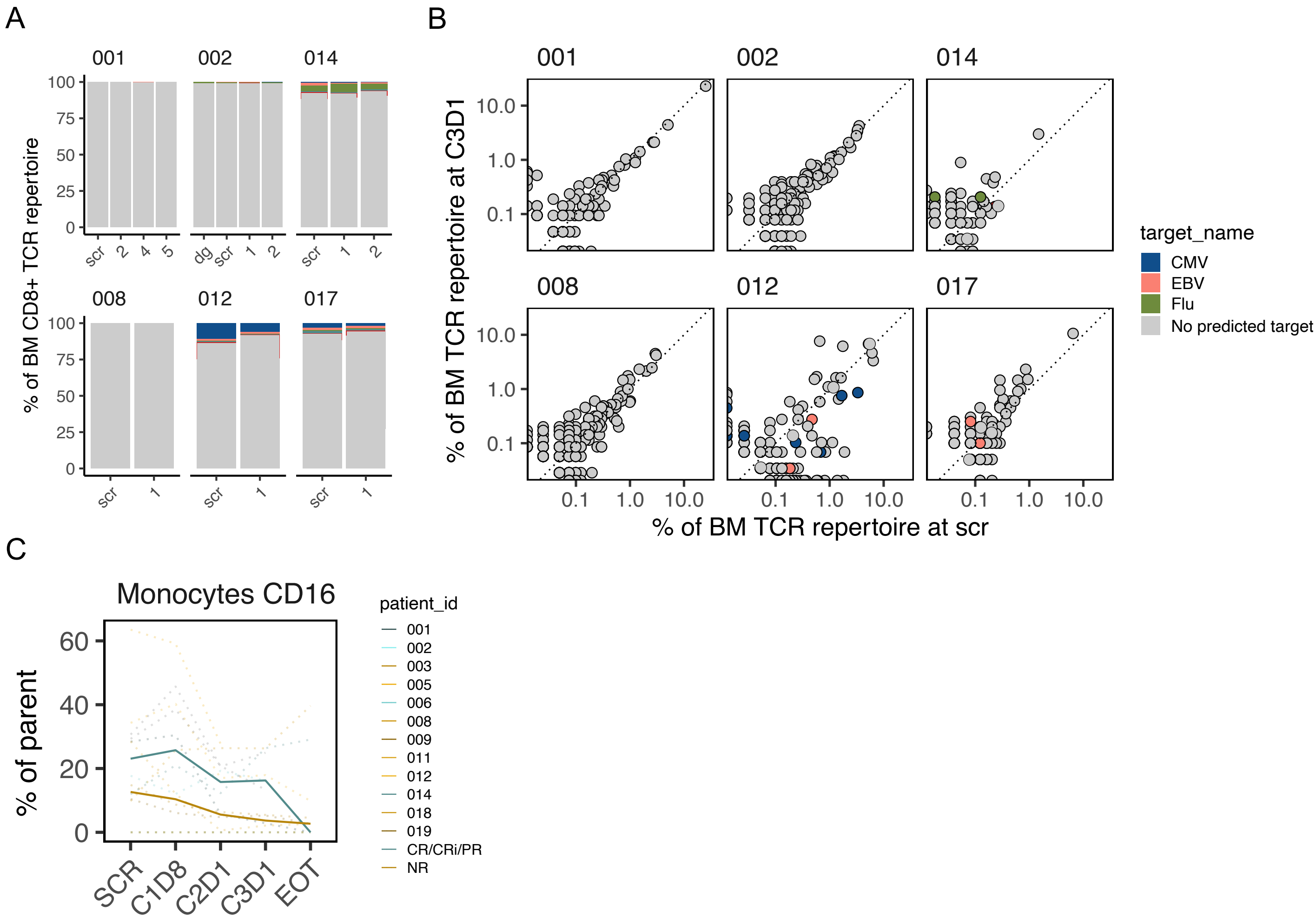

**Supplementary Figure 11: TCRGP prediction of targets**

**A)** Proportion of CD8+ T cells carrying TCRs predicted to recognize endemic viruses with TCRGP.
**B)** TCR clone proportions between pre-treatment and C3D1 timepoints, where TCRGP predictions have been highlighted.
**C)** The proportion of CD16+ monocytes out of total CD45+ cells in peripheral blood across patients. The enforced lines show the median across responders and non-responders.

Supplementary Figure 12

A

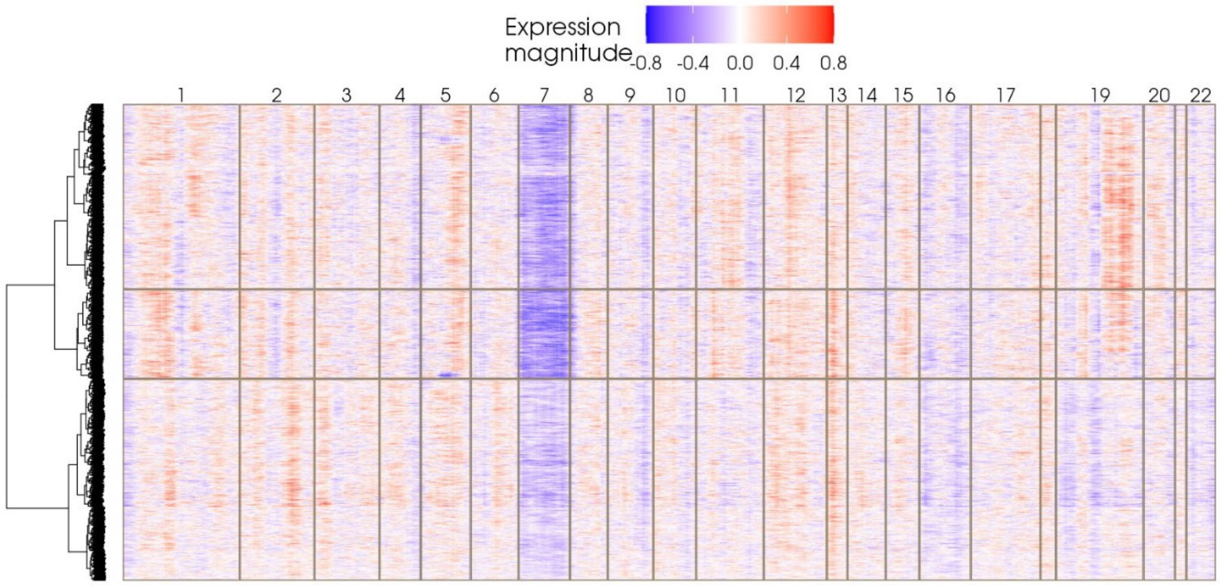

**Supplementary Figure 12: Patient case 001**

**A)** Heatmap of the CNV analysis results with Numbat showing the expression magnitude in individual chromosomes in the patient 001 relapse sample.

Supplementary Figure 13

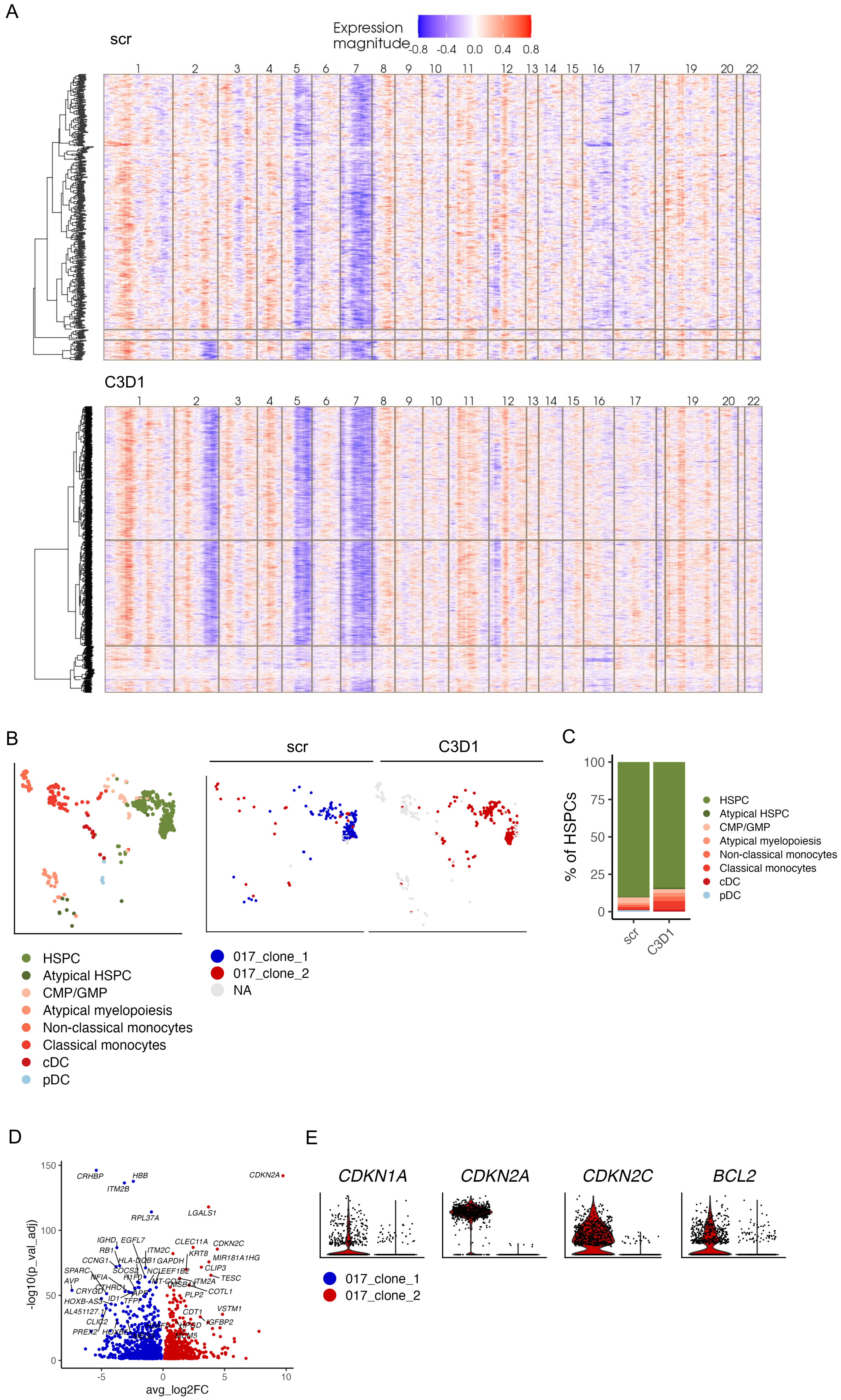

**Supplementary Figure 13: Patient case 017**

**A)** Heatmap of the CNV analysis results with Numbat showing the expression magnitude in individual chromosomes in the patient 017 screening (top) and C3D1 (bottom) samples.
**B)** UMAP of the MDS cells from patient 017 in total (left) and across different time points (right).
**C)** Proportion of different phenotypesfrom patient 017 across different time points.
**D)** Differentially expressed genes between clone 2 and clone 1 in patient 017.
**E)** Violin plot showing the most dysregulated genes between clones 2 and 1 in patient 017.

**Supplementary Note 1: Exceptional responder patient 001**

A female in their 70s, previously treated for breast cancer in 2006 with postoperative radiotherapy (50 Gy), and diagnosed with CD4+ T-large granular lymphocytic leukemia (T-LGLL) in 2012 after slight neutropenia that required no treatment. Subsequent progression to secondary AML was observed in 2017, characterized by mutations in TET2, SRSF2, RUNX1, and deletion 7, along with a variant of uncertain significance (VUS) in GATA2. Conventional chemotherapy was contraindicated due to the disease's nature and patient preferences, and thus the patient was enrolled in our clinical trial.

During cycle 1, the patient experienced mild thrombocytopenia and elevated ALAT values that were successfully treated with cortisone therapy. Simultaneously, the leukocytes increased and the C1D22 bone marrow showed progressive disease. The treatment was discontinued and the patient referred to palliative care.

However, what would have been C2D17 all the blood counts normalized spontaneously, and C2D18 bone marrow showed CR with slight MRD positivity (FISH: 2/777 interphases). The patient was enrolled again to the therapy and C3D1 was started on 21.11.2017. C6D1 bone marrow showed MRD negativity. After two years, C19D23 BM sample showed MRD positivity (FISH: 2/1000 interphases), but C20D6 was MRD negative again. However, in C24D2 showed a morphological relapse, and MRD positivity (FISH 2.1%). The bone marrow showed an increase in lymphocytes and plasma cells (5-10%). The bone marrow did not show any more T-LGLL clone, but an increase in NK cells. The treatment was continued but eventually stopped 31.1.20 due to PD.
